# The Modest Impact of Weather and Air Pollution on COVID-19 Transmission

**DOI:** 10.1101/2020.05.05.20092627

**Authors:** Ran Xu, Hazhir Rahmandad, Marichi Gupta, Catherine DiGennaro, Navid Ghaffarzadegan, Heresh Amini, Mohammad S. Jalali

## Abstract

Understanding how environmental factors impact COVID-19 transmission informs global containment efforts. We studied the relative risk of COVID-19 due to weather and ambient air pollution. We estimated the daily reproduction number at 3,739 global locations, controlling for the delay between infection and detection, associating those with local weather conditions and ambient air pollution. Controlling for location-specific fixed effects and local policies, we found a negative relationship between the estimated reproduction number and temperatures above 25°C, a U-shaped relationship with outdoor ultraviolet exposure, and weaker positive associations with air pressure, wind speed, precipitation, diurnal temperature, SO_2_ and ozone. We projected the relative risk of COVID-19 transmission due to environmental factors in 1,072 global cities. Our projections suggest warmer temperature and moderate outdoor ultraviolet exposure may offer a modest reduction in transmission; however, upcoming changes in weather alone will not be enough to fully contain the transmission of COVID-19.

## Introduction

The COVID-19 pandemic has significantly challenged the global community. High-stakes policy decisions depend on how environmental factors impact the transmission of the disease (*1*). Given that many related viral infections such as seasonal flu (*2*), MERS (*3–5*), and SARS (*6*) show notable seasonality, one may expect the transmission of SARS-CoV-2 virus to be similarly dependent on weather. Earlier works indicate that temperature (*7*), humidity (*7–9*), air pressure, ultraviolet light exposure, and precipitation may impact the spread of COVID-19 by changing the virus survival times on surfaces and in droplets (*10–12*), moderating the distance virions may travel through air (*12*), changing host susceptibility, and impacting individual activity patterns and immune systems (*7, 8, 10–12*). A few other studies suggest air pollutants may act as vectors for the virus or impact the immune system (*13, 14*).

Yet, there is limited agreement on the shape and magnitude of those relationships. While studies find correlations between pandemic severity and variations in temperature (*9, 15–22*), relative and absolute humidity (*9, 16–24*), ultraviolet light (*19*), wind speed (*18, 21*), visibility, and precipitation (*17*), others (*19, 25–27*) indicate weaker, inconsistent, or no relationships. A recent review finds inconclusive evidence for the role of weather in COVID-19 transmission (*1*) and others caution against interpreting weather as a key driver due to this uncertainty (*28*).

The explanation for these inconclusive results is unclear. Estimates that are based on datasets focused only on China or the United States (*17, 19, 20, 22–24, 29, 30*) may be too narrow. Others have studied only a subset of meteorological measures (*9, 16, 22–24, 27, 29*), complicating comparisons. Most studies have not controlled for other important factors such as varying government and public responses, population density, and cultural practices (*9, 17–19, 24, 30, 31*). The delay between infection and official recording of cases is a particularly understudied factor. Failure to correct for these delays, estimated to be approximately 10 days (*32, 33*), confounds attempts to associate daily weather conditions with recorded new cases and may partially explain the inconsistent and inconclusive findings to date.

Here, we assemble one of the most comprehensive datasets of the global spread of COVID-19 pandemic through late April 2020, spanning more than 3,700 locations around the world. We validate and apply a statistical method to estimate the daily reproduction number in each location. Controlling for location-specific differences in population density, cultural practices, socio-economic differences, public transportation, nutrition, age distribution, and time-variant responses in each location (e.g., physical distancing, quarantine, lock-down, public space closures), we estimate the association of weather and air pollutants with the reproductive number of COVID-19 and provide year-round, global projections.

## Data and Methods

### Data

Our dataset includes infection data for 3,739 distinct locations, spanning the beginning of the epidemic (December 12, 2019) to April 22, 2020. We augment the data reported by the Johns Hopkins Center for Systems Science (*34*) with data reported by the Chinese Center for Disease Control and Prevention, Provincial Health Commissions in China, and Iran’s state-level reports. We assemble disaggregate data on the spread of COVID-19 in Australia (8 states), Canada (10 states), China (34 province-level administrative units and 301 individual cities), Iran (31 provinces), and the United States (3,144 counties and 5 territories) and use country-level aggregates for the remaining 206 locations.

We compile weather data from archival databases (World Weather Online, and OpenWeather Ltd.), and air pollution data from the European Centre for Medium-Range Weather Forecasts. For country-level locations that include cities with populations 500,000 or greater, weather and pollution data were first gathered for each city and then averaged over all cities weighted by their population into country-level measures. For U.S. counties, Canadian and Australian provinces, and any remaining countries, we use the weather and air pollution data for the coordinate of the centroid of that location. We obtain daily data for minimum and maximum temperature, humidity, precipitation, snowfall, moon illumination, sunlight hours, ultraviolet index, cloud cover, wind speed and direction, pressure data, as well as air pollutants including ozone (O_3_), nitrogen dioxide (NO_2_), sulfur dioxide (SO_2_), and particulate matter with aerodynamic diameter ≤ 2.5 μm (PM_2.5_). We used population density data from Demographia (Cox., W, Demographia, The Public Purpose), the United States Census (U.S. Census Bureau), the Iran Statistical Centre, the United Nation’s Projections, City Population (citypopulation.de), and official published estimates for countries not covered by these sources (United Nations).

### Methods

#### Estimation of the Reproduction Number

A critical parameter in understanding the spread of an epidemic is the effective reproduction number, *R_e_*, the expected number of secondary cases generated by an index patient. An epidemic grows when *R_e_* is above 1 and will die out once *R_e_* stays below 1. Reproduction number can be approximated (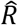) based on the number of new infections (*I_M_*) per currently infected individual, multiplied by the duration of illness (*τ*). Actual new infections on any day (*I_N_*) are not directly observable but an unbiased estimator, *Î_N_*, can be used to estimate 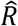 (equation 1). Data on measured daily infections (*I_M_*) lag actual new infections by both the incubation period and the delay between the onset of symptoms and testing and recording of a case. We use published measures to quantify the distribution of both the incubation period and onset-to-detection delays (averaged between 5 to 6 days (*20, 32, 33, 35*) and between 4 to 6 days (*32, 33*), respectively). Together these shape the overall *detection delay*. Given the variance in detection delay, a simple shift of measured infection by the mean delay (about 10 days) offers an unreliable estimate of true infections (see Section S3 and S5.2.2.2 in Supplementary Document). We therefore develop an algorithm to find the most likely actual daily new infections (*Î_N_*) based on the observed measured infections (*I_M_*) and the detection delay distribution (see S3). We then use *Î_N_* to estimate the reproduction number:

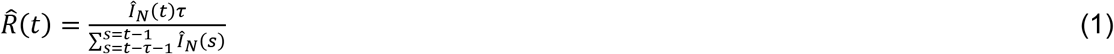

We use the daily 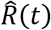 as our dependent variable. The estimate of 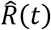 is robust to the existence of asymptomatic cases, under-reporting, and changing test coverage as long as the changes in test coverage are uncorrelated with weather conditions 10 days ago (see S3 and S5.2.2.3 for details). We use a delay of *τ*=20 days from exposure to resolution; results are robust to other durations of illness (see S4.1). For each location, we only include days with *Î_N_* values above one. Reliability of early *Î_N_* values for each location is affected by irregularities in early testing. Moreover, an unbiased estimate for 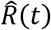 requires *τ* days of prior new infection estimates. Thus, to ensure robustness we exclude the first 20 days after *Î_N_* reaches one in each location (S4 discusses robustness to these criteria and the exclusion of outliers).

#### Controls for estimating 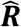

The reproduction number for COVID-19 primarily varies due to location-specific factors, from population density, cultural practices, and public transportation use to nutrition, age distribution, and genetic profile, among others. We control for these and other unobserved factors using location fixed effects. Moreover, school closures, gathering bans, distancing, and other behavioral responses reduce *R_e_* over time. Reproduction number may also increase if endogenous epidemic takes off later (e.g. when contact tracing is overwhelmed). We account for such changes by estimating a location-specific time trend in *R_e_* and assess sensitivity to nonlinear trend controls in S4. We also separately control for day of the week.

#### Independent Predictors

Prior studies (*9, 20*) suggest *R_e_* may depend on various meteorological and air pollution factors through at least three pathways. First, the survival of the virus on surfaces and the spread of droplets and particles containing the virus may be impacted by temperature, UV, humidity, wind, and particulate matter (*12, 13*). Second, human host susceptibility may be impacted due to factors moderating immune response (e.g., impact of UV on serum Vitamin D (*19*)) as well as respiratory tract susceptibility to virus (e.g., temperature, humidity, and pollutants (*9, 14*)). Finally, the behavior of human hosts (e.g., interacting indoors and outdoors) is likely affected by multiple factors, ranging from temperature and precipitation to air pollutants, UV index, and humidity (*27*). Our data do not allow us to tease out these distinct pathways explicitly. Instead, we include the following potential contributing factors, focusing on their direct effects in the main specification and discussing interactions in the Supplementary Document: temperature (mean and diurnal temperature (difference between maximum and minimum daily temperature), relative humidity, pressure, precipitation, average wind speed, and ultraviolet exposure (*UV* Index). We included O_3_ and SO_2_, as air pollutants. We also explored a few interactions among these variables as well as the inclusion of other environmental variables including absolute humidity, number of sun hours received, snowfall, moon illumination, NO_2_, and PM_2.5_ and report those results in S4.

#### Statistical Specification and Validation

Given the large variations in 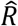 estimated in this method, we use a log transformation of 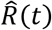 and linear models to predict 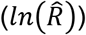. We designed and validated our statistical model for estimating 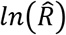 by testing its ability to identify true parameters in synthetic data. Specifically, we built a stochastic simulation model of the COVID-19 epidemic, generated synthetic infection data using historical weather inputs and presumed impact functions, and designed a statistical model that reliably found those presumed effects under an ensemble of simulated epidemics with different basic reproduction numbers, weather effects, population sizes, and test coverage, among others. We found that: a) given actual infections (*I_N_*), our method identifies the presumed functional form relating weather to transmission rate; b) estimates become conservative (between null and true effects) when true infections are inferred rather than known; c) our method for inferring infections offers significantly better results than a simple shifting of official counts (see Sections S3 and S5.2.2.2).

Separately, to independently validate the resulting statistical method, authors NG and MG created a realistic individual-based model of disease transmission and used that to generate a separate test dataset with synthetic epidemics. Three scenarios were created using actual temperatures from a sample of 100 regions and three different functions for the temperature effect. Then author RX, who was blinded to the true functions in this synthetic dataset, successfully estimated the correct qualitative shape of those functions using our method (Section S5.3).

Building upon these findings, our main specification excludes days with *Î_N_* <1 and the first 20 days after *Î_N_* exceeds one for the first time. The model includes location-specific fixed effects and trends and includes the following effects: a linear spline for the effect of average temperature on transmission (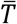), with the knot at 25°C (see Section S4.5 for alternative knot values), diurnal temperature (Δ*T*), air pressure (*P*), relative humidity (*H*), linear and quadratic effects of Ultraviolet Index (*UV*), log precipitation (*C*), log wind speed (*W*), log *O*_3_, and log *SO*_2_:

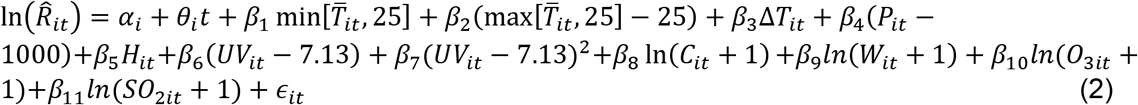

#### Projection

We project the impact of weather and air pollution on the relative risk of transmission for all locations in our sample as well as 1072 major urban areas (population > 0.5 million) constituting about 30% of world’s population. A summary of results is provided in the paper and an interactive online platform offers comprehensive projections.

## Results

### Estimated impact of weather and air pollution on transmission rate

Table 1 reports our main results. The model explains roughly three quarters of the variance in 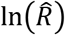 values (R^2^=.740), much of which due to fixed effects (39.2% of variance) and trends (34.6 % of variance). Average initial 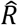 is 1.98 (IQR: 0.88 to 2.49) 20 days after the first estimated case with much variation across locations (Figure 1-A). Initial 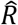 is negatively correlated with epidemic start time and positively with population density. Most locations show rapid reductions in reproduction number over time that capture the impact of policies and behavioral changes that reduce contacts. On average, 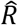 falls 5.8% (IQR: −1.7% to 8.7%) per day but with notable variability across locations (Figure 1-A) partly explained by locations with higher initial 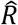 having faster subsequent reduction. For example, after excluding the first 20 days of estimated infections, New York City shows an initial 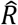 of 5.07, followed by a 7.8% daily reduction (Figure 1-B).

**Table 1.** Impact of weather on COVID-19. Outcome variable: 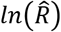

| Weather variables | Mean (SD) | Coefficient (95% CI) | p-value | Standardized coefficient* |
| --- | --- | --- | --- | --- |
| Wind speed (log of Km/Hour) | 2.552 (.444) | .0323 (.0079 .0567) | .01 | .0144 |
| Precipitation (log of Millimeters) | .785 (1.022) | .0265 (.0168 .0361) | <.001 | .0271 |
| Air pressure (millibars) | 1015.47 (6.08) | .0022 (.0005 .0039) | .013 | .0132 |
| Humidity (Percent) | 66.946 (15.075) | -.0006 (-.0015 .0003) | .179 | -.0091 |
| Mean temperature below 25 (Celsius) | 11.351 (7.089) | -.004 (-.0066 -.0014) | .003 | -.031 |
| Mean temperature above 25 (Celsius) | 27.761 (2.204) | -.0377 (-.0559 -.0194) | <.001 | -.0391 |
| Ultraviolet index (25 milliwatts/m²) | 7.13 (2.824) | .0089 (-.0083 .0260) | .312 | .0250 |
| Ultraviolet index^2 |  | .0053 (.0025 .0081) | <.001 | .0702 |
| Diurnal temperature (Celsius) | 8.757 (3.315) | .0042 (.0012 .0072) | .006 | .0139 |
| Ozone (log of ppbv) | 3.289 (.667) | .0349 (.0161 .0537) | <.001 | .0232 |
| Sulfur Dioxide (log of ppbv) | 1.139 (.936) | .0301 (.0118 .0485) | .001 | .0282 |
|  | Mean (SD) over i | Mean of Standard Error Across Locations |  |  |
| Fixed Effects ($\ln(\hat{R}_i(0)) = \alpha_i$) | .377 (.767) | .519 | | |
| Trends ($\theta_i$) | -.060 (.157) | .107 | | |
| N=19,221; R²=.740 |  |  |  |  |

\*Standardized coefficients were obtained by first standardizing all of the weather variables (mean=0, SD=1) and then re-running the analysis with our main model specification.

**Figure 1.**
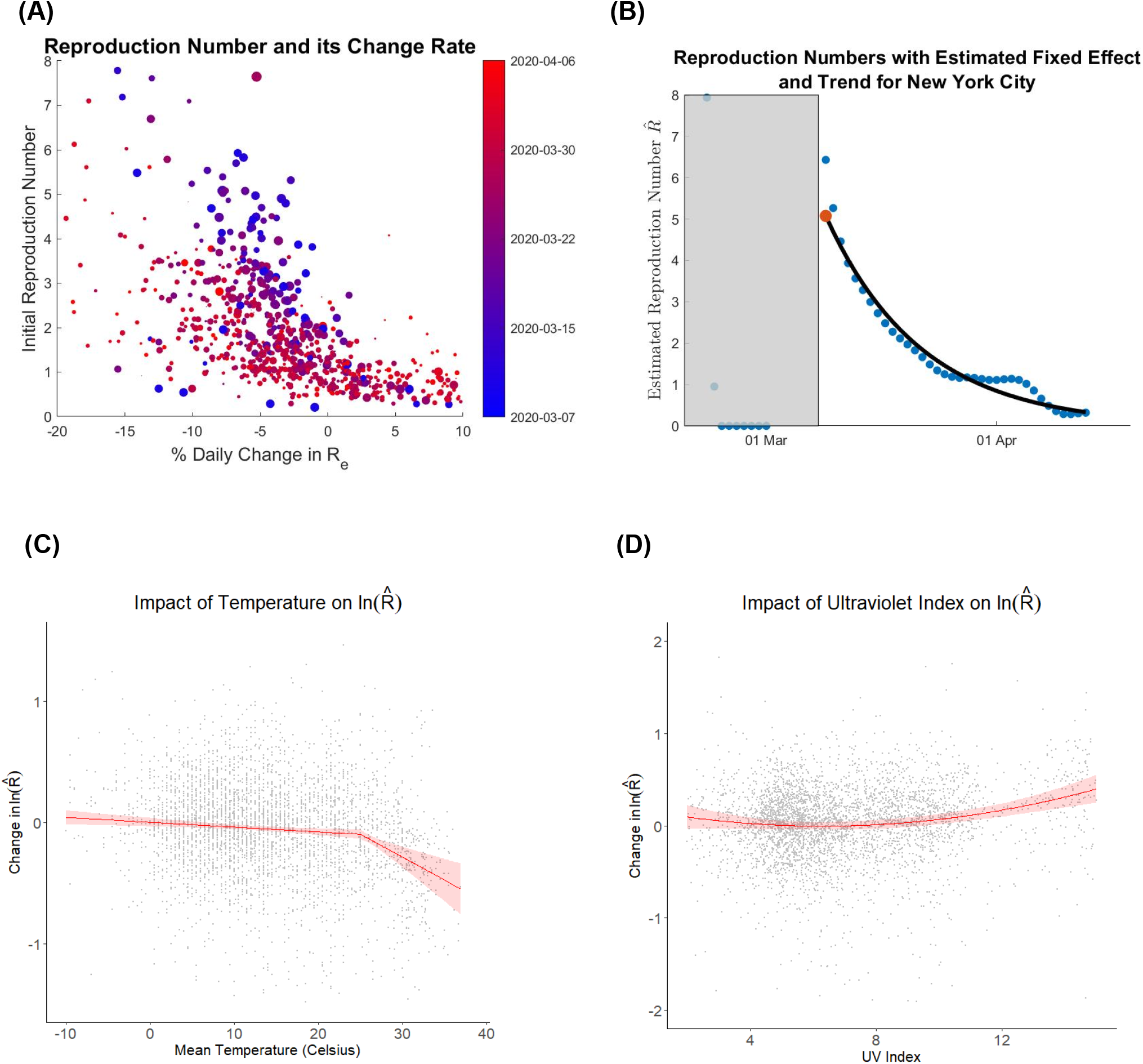
Summary of results. A) Scatter diagram of Initial Reproduction Number (Y-axis) and Daily % Change (X-axis) across different locations, color coded for date of local epidemic start. Circles’ sizes scale with population density in a location. B) Estimated daily 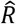 values for New York City (blue dots; 20 days in gray area excluded); the estimated initial 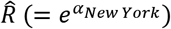 (Red Dot) and trend (Black line). C) Relationship between temperature and reproduction number (β_1_ and β_2_) with its uncertainty. D) Relationship between UV index and reproduction number. Panels C and D include (downsampled) data and control for other factors as in Eq. 2.

Even after controlling for these factors, mean temperature, Ultraviolet index, diurnal temperature, air pressure, wind speed, precipitation, *O*_3_, and *SO*_2_ are significantly associated with transmission (Table 1). We found a robust effect of mean daily temperature, which is best characterized within two regimes, below and above 25°C (Figure 1-C). Temperatures higher than 25°C were associated with lower transmission rates (by 3.7% (CI: 1.9-5.4%) per additional degree) while those below that threshold had a smaller impact (0.4% (0.14-0.66) reduction per degree).

UV has a robust U-shaped effect on the reproduction number, with a minimum around 6.3 (Figure 1-D). At a low/moderate UV of 3, a unit higher UV decreased 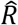 by 3.5% (0.4-6.4%). At a high UV of 10, a unit higher UV increased 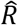 by 4% (1.8-6.3%). While less robust across specifications (see Section S4), we also find weak/moderate and statistically significant positive effects of diurnal temperature, air pressure, wind speed, precipitation, O_3_, and SO_2_. A one standard deviation increase in each increases 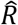 by 1.4% (0.4-2.4%) for diurnal temperature, 1.3% (0.3-2.4%) for air pressure, 1.4% (0.4-2.5%) for log-transformed wind speed, 2.7% (1.7-3.8%) for log-transformed precipitation, 2.4% (1.1-3.6%) for log-transformed O3, and 2.9% (1.1-4.6%) for log-transformed SO2.

We also find a few interactions among these predictors may be relevant in determining transmission rates (Not included in main model; See 4.7 for details): the quadratic effect of ultraviolet index precipitation is dampened with the increase in precipitation; the negative effect of mean temperature above 25 °C is attenuated with higher SO_2_ levels; and there may be a positive effect of PM_2.5_ which is attenuated with increased air pressure. Finally, we found a significant reduction in transmission rate associated with higher moon visibility, but lacking a theoretical explanation for the effect, we did not include it in the main specification.

Overall, the association of various weather and air pollution variables with COVID-19 transmission is large enough to be relevant to assessing the risk of across locations and seasons. Variations in the reproduction number associated with the combined set of predictors in our estimation dataset showed a ratio of 1.24 between the 95^th^ and 5^th^ percentile despite the sample largely coming from late winter/early spring. Given that the typical reproduction number estimated for COVID-19 is in the range of 2 to 3 (*36, 37*), estimated weather effects alone may not provide a path to containing the epidemic in most locations, but could notably impact the relative transmission rates.

### Robustness

Validation of our statistical method using synthetic data (S5.2) showed that: (a) our results are robust to under-reporting as well as changes in test coverage; (b) our method can identify the correct sign and shape for the impact of environmental variables; and (c) those estimates are potentially conservative (i.e., smaller than the true impacts). The conservatism is due to two factors. First, unavoidable errors in estimating daily infections from lagged official data lead to imperfect matching of independent variables and true infection rates, weakening any estimated relationship. Second, fixed effects further weaken the signal used for estimation: If a region has a lower/higher baseline reproduction number due to weather factors, that effect is absorbed in the fixed effects and will not impact the estimates for relevant factors.

We also conducted eight empirical tests to assess the robustness of our findings (See S4). First, our results do not change with the use of different illness durations to calculate 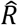. Second, our main findings are robust to excluding extreme values of the dependent variables, the last few days of data, only using the USA sample, and the inclusion of location-specific quadratic trend or time fixed effects. Third, our results are largely insensitive to different exclusion criteria for initial periods of transmission per location. Fourth, when independent variables in each location are permuted and shifted in a placebo test, no effects remain, showing that results are not artifact of statistical method. Fifth, using different knots for the spline effect of temperature shows 25°C best separates the effect into two distinct slopes. Sixth, the estimated U-shaped effect of UV does not change when observations with high UV index are excluded. Seventh, we explored more interaction terms, additional weather variables (e.g. absolute humidity, nitrogen dioxide and PM_2.5_), none of which change the main results. Finally, overall projections of how weather and air pollution impact transmission rates using various specifications and on independent samples are consistent with our main specification.

### Projections

Our results are associative and extrapolating out-of-sample includes unknown risks. With that caveat in mind, one can calculate the contribution of weather and air pollution to expected transmission for any vector of weather and air pollution based on Table 1 results. We defined “Relative COVID-19 Risk Due to Weather and air pollution” (CRW) as the relative predicted risk of each weather and air pollution vector relative to the 95^th^ percentile of predicted risk in our estimation sample (1.476). The choice of this reference point is somewhat arbitrary but makes a value of 1 a rather high-risk level. A CRW of 0.5 reflects a 50% reduction in the estimated reproduction number compared to this (high-risk) reference. Formally:

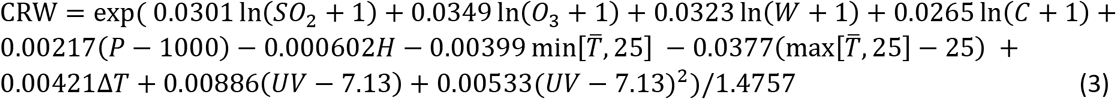

These scores do not reveal the actual value of *R_e_* as that value is contingent on location-specific factors and policies for which we have no data outside the estimation sample. Rather, CRW scores inform relative risks due to weather and air pollution (i.e., assuming all else equal) across locations or within a location over time.

Figure 2 provides a visual summary of global CRW scores, averaged over the first half of July 2020. The color-coded scores suggest much variation in the expected risk of COVID-19 transmission across locations, with increased risks due to both low temperature (some regions in southern hemisphere) and very high UV indexes (some locations in central America). Section S6 provides additional snapshots of global CRW scores at different times of the year, and the website (projects.iq.harvard.edu/covid19) offers week-by-week risk measures year-round.

**Figure 2.**
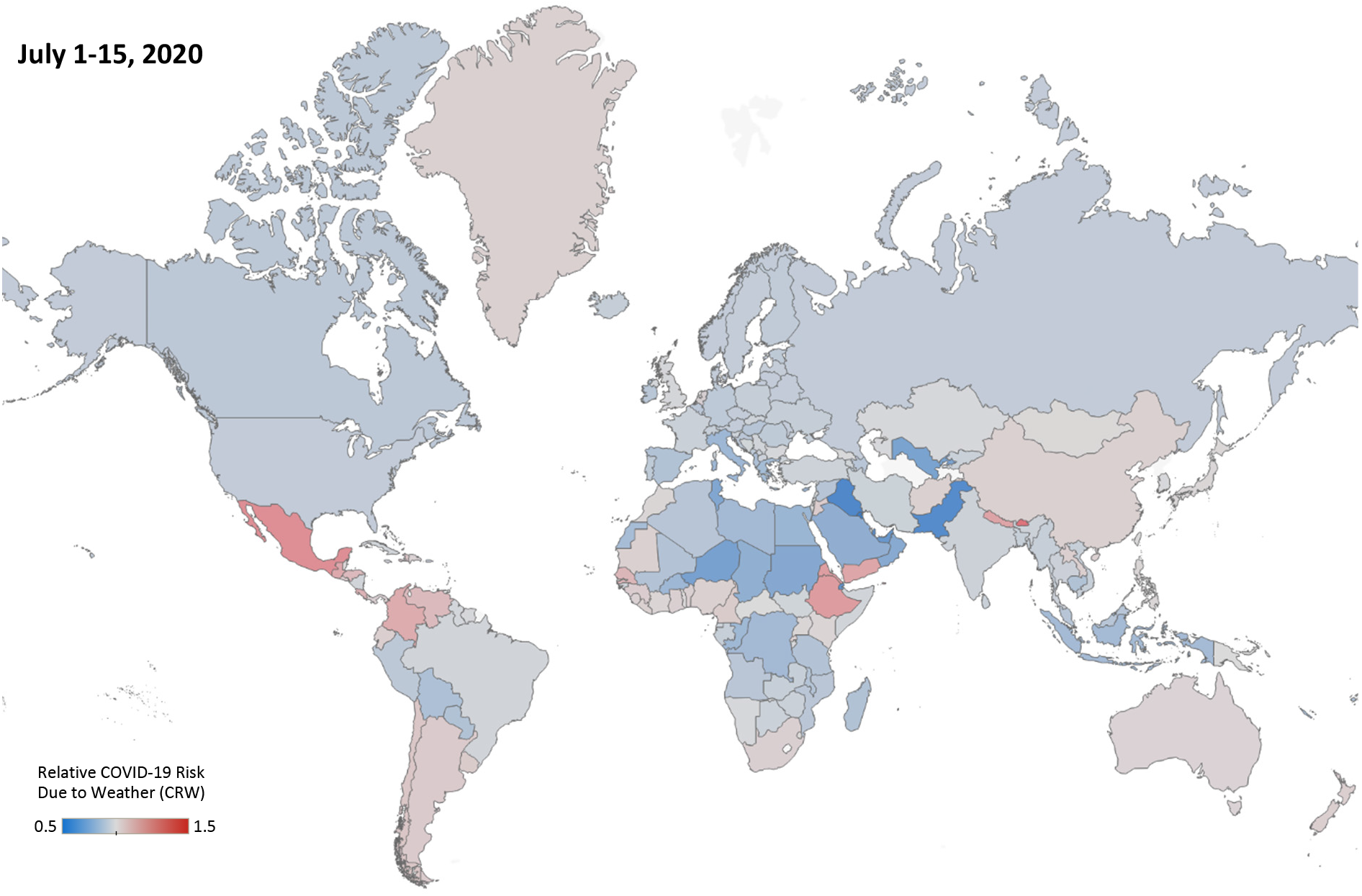
Relative COVID-19 Risk Due to Weather and air pollution (CRW) for different regions of the world, averaged over the first half of July 2020.

Figure 3 shows CRW projections for five major cities in each of the four regions of America (panel A), Europe (B), Africa and Oceania (C), and Asia (D). These projections use prior year weather and air pollution from 2019, averaged over a 15-day moving window, for 2020-2021 dates; as such, they include historical noise despite the 15-day averaging. Many large cities go through periods of higher and lower risk during the year. We cannot associate these risks with absolute reproduction numbers, and our estimates are likely conservative. Nevertheless, assuming typical basic reproduction rates (e.g., 2-3), weather factors will not bring the reproduction number below 1. For example, in New York City, with estimated 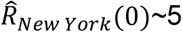, the impact of weather may lead to a 30% variation in the reproduction number (i.e., the 4 to 6 range), requiring significant social distancing policies to enable containment regardless of weather. The website projects.iq.harvard.edu/covid19 provides these projections for the 1,072 largest global cities.

**Figure 3.**
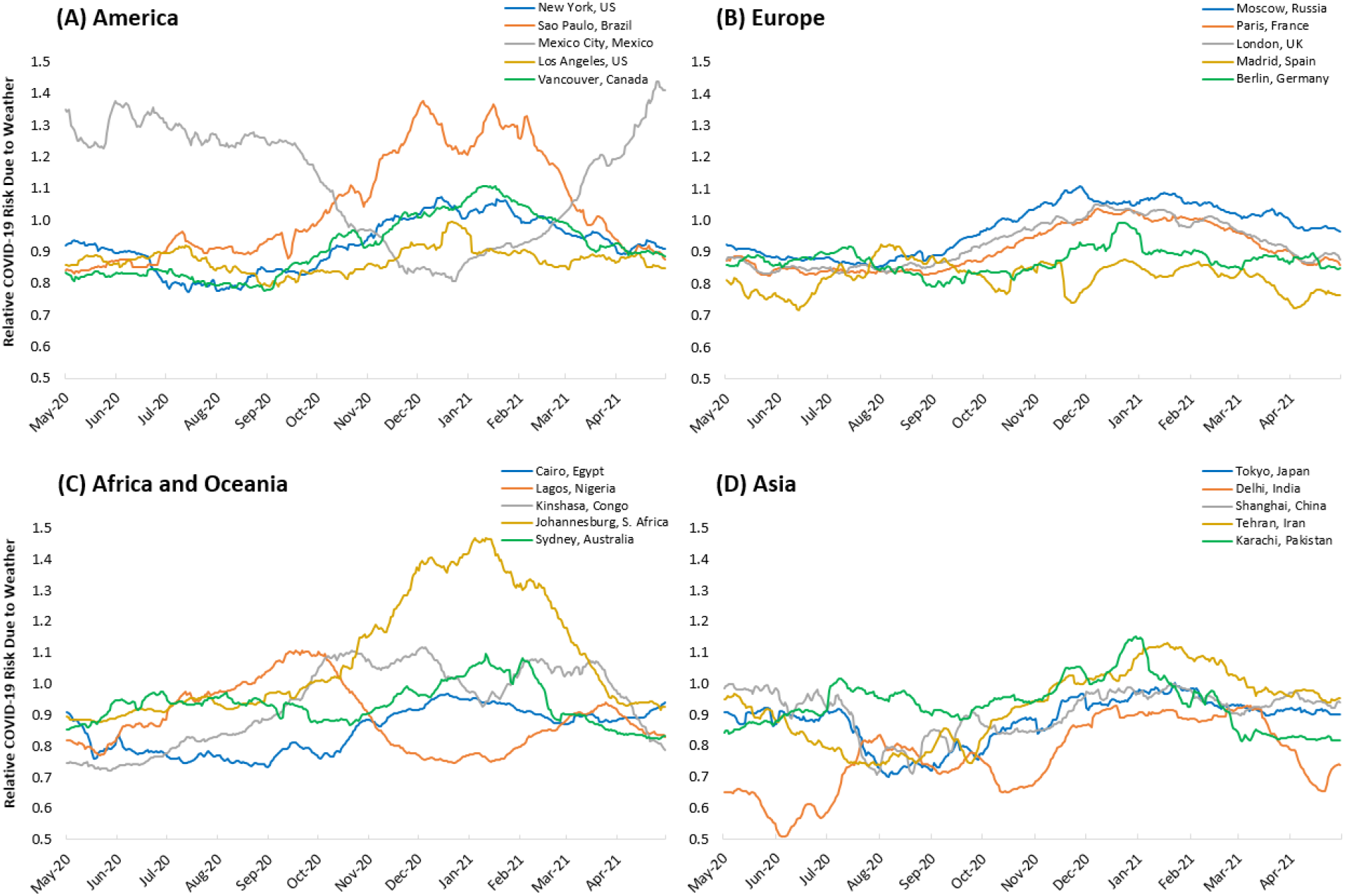
CRW measures over the year for major cities around the world.

## Discussion

This work combines one of the most comprehensive datasets of COVID-19 transmission with weather and air pollution data across the world to estimate the association of various environmental variables with the spread of COVID-19.

We find a strong association between temperatures above 25°C and reduced transmission rates, and a weaker effect below 25°C. These suggest many temperate zones with high population density may face larger risks in winter, while some warmer areas of the world may experience slower transmission rates in general. The U-shaped relationship between UV index and transmission may help more temperate regions during summer, but higher risks in equatorial regions with very high UV exposure.

Most of the effects we found are consistent with theoretical mechanisms thought to link environmental factors to transmission: the negative temperature effect on transmission, boosted at higher temperatures, is consistent with virus survival rates in experimental work (*12*); the positive effects of wind and precipitation could result from people spending more time indoors where transmission is more likely than outdoors; and the impact of air pollutants may be related to increased susceptibility in more polluted environments (*14*). Nevertheless, we are mindful that our study design and data cannot tease out such mechanisms empirically. For example, we hypothesized a UV exposure to reduce transmission (due to both stimulating vitamin D production and UV’s disinfecting effects). Our estimates have the expected sign in the low ranges of UV, but also reveal an unexpected increase in the high ranges of UV. The latter effect may be due to a shift of social interactions into higher risk, indoor, settings when UV levels are very high; but we cannot test such explanations here.

Methodologically, we show that accounting for the distribution of the delay between infection and detection is important. Many prior studies did not fully account for this delay, or its distribution, which may partially explain inconsistent prior results. We also showed (in S5.2.2) that our methods and results are robust to significant under-counting of cases in official data, as well as to changes in test coverage over time, both major concerns in using official case data.

Nevertheless, estimates of *R_e_* are imperfect, leading to conservative overall estimates for various effects, a fact that should be noted in using our projections. Other limitations include: the lack of reliable transmission data in some regions of the world; oversampling from U.S. locations; limited data with high temperature and UV in our estimation sample, which reduce confidence for projections when either is very high; use of last year’s weather data to project next year’s outcomes; and use of correlational evidence to inform out-of-sample projections.

Despite these limitations, consistent results using various conservative specifications and placebo and validation tests provide promising indications of the true impacts of weather conditions on transmission. The estimated impacts suggest summer may offer partial relief to some regions of the world. However, given a highly susceptible population, the estimated impact of summer weather on transmission risk is not large enough in most places to quell the epidemic in 2020, indicating that policymakers and the public should remain vigilant in their responses to the pandemic. In fact, much of the variation in reproduction number in our sample is explained by location-specific fixed effects and responses, not weather; and most regions that can expect reduced risk in summer will face increased risks in the fall. Ultimately, weather more likely plays a secondary role in the control of the pandemic.

## Data Availability

All data and analysis codes are available on the project website: https://projects.iq.harvard.edu/covid19

## Data, codes, and online simulator

https://projects.iq.harvard.edu/covid19

## Acknowledgements

We thank John Sterman, Richard Larson, Goodarz Danaei, Neil Rubin, Robert Andrew Brown, Mark Shumate, Pari Pandharipande, and Saeid Shahraz who provided feedback and suggestions on earlier versions of this manuscript. We thank Cynthia Dong for insights on weather data collection, Peiyi Li, Hongjin Xu, and Wenhan Dai who assisted us in collecting and evaluating data from China, and Maedeh Moghadam who helped us with assembling data from Iran.

## Online Supplementary Document for

### 1. Data and replication instructions

All code and data for this research are available at https://github.com/marichig/weather-conditions-COVID19.

Case and coordinate data were first taken from JHU’s published case reports, available at https://github.com/CSSEGISandData/COVID-19, which covered all locations (counties) in the United States and 258 of the 590 locations from outside the U.S., including breakdowns for Canada into 10 states/territories, and Australia into 8 states. The remaining locations include 301 Chinese cities and 31 Iranian states; for these, case and coordinate data were taken from the Chinese Center for Disease Control and Prevention, Provincial Health Commissions in China, and Iran’s state level reports.

Some locations included in the U.S. case reporting data were dropped from the main analysis. Namely:

- Cases from the cruise ships Diamond Princess, Grand Princess, and MS Zaandam were discarded.
- Cases labelled as “Out of [State]” or “Unassigned, [State]” were discarded.
- Cases from Michigan Department of Corrections and Federal Correctional Institute, Michigan were dropped since they reflect unique spread dynamics and carried no coordinate data.
- Cases attributed to the Utah Local Health Departments (Bear River, Central Utah, Southeast Utah, Southwest Utah, TriCounty, and Weber-Morgan) were discarded; as of 4/22/2020, only 291 cases were reported from these sources compared to 3154 from all Utah counties. These health departments span several counties and reporting from them only began on 4/19/2020.

Errors in the reported coordinate data were also identified and resolved manually. (For instance, Congo-Brazzaville was reported to have the same coordinates as Congo-Kinshasa.) With this coordinate data, weather data is collected primarily through World Weather Online (WWO), which provides an API for data collection – the Python “wwo-hist” package <https://pypi.org/project/wwo-hist/> was used to access this API. Historical weather data were collected for each day between 1/23/2019 thru 4/22/2020, with data from 2019 being used for future projection. Pollution data are collected from the European Centre for Medium-Range Weather Forecasts (ECMWF)’s CAMS-Near Real Time service from 1/1/2019 – 4/22/2020, with solely 2019 data used for projection, since 2020 data is not representative due to disruption of human activity from the pandemic.

The following weather variables were collected: maximum daily temperature (degrees Celsius (°C)), minimum daily temperature (°C), average daily temperature (°C), precipitation (millimeters), humidity (percentage (%)), atmospheric pressure (millibars), windspeed (kilometers per hour (km/h)), sun hours (i.e., hours of sunshine received), total snowfall, (centimeters) cloud cover (percentage) ultraviolet (UV) index (measured within one hour of noon local time), moon illumination (%) (i.e., percentage of moon face lit by the sun), local sunrise and sunset time; local moonrise and moonset time; dew point (°C), “Feels Like” (°C), wind chill (°C) wind gust (i.e., peak instantaneous speed) (km/h), visibility (kilometers), and wind direction degree, clockwise degrees from due north. The pollutant variables collected were ozone (parts per billion volume (ppb (v)), nitrogen dioxide (ppb (v)), sulfur dioxide (ppb (v)), and particulate matter of diameter less than 2.5 micrometers, micrograms per cubic meter. Descriptions of the weather variables are available at <https://www.worldweatheronline.com/developer/api/docs/historical-weather-api.aspx>. The ultraviolet (UV) index data were not consistently reported from WWO, and were instead gathered using OpenWeatherMap <https://openweathermap.org/> and the Python “pyowm” package <https://pypi.org/project/pyowm/>.

We interpolated over any missing entries in the temperature, UV, and pollution data provided. The reported temperature data were missing for most (but not all) locations for a handful of days: 9/15-17/2019, 10/22/2019, 11/27/2019, and 12/15/2019, which were then interpolated using five-day moving averages. UV data were missing for less than 0.1% of date-location pairs, with the main gaps occurring on 6/2/2019, 8/13/2019, 12/2/2019, 2/18/2020, and 2/21/2020, which were interpolated using three-day moving averages. The averaging of temperature and UV data should not impact the analysis given that most of the above dates fall outside the pandemic’s date-range. Furthermore, across all pollution variables, less than 0.1% of date-location pairs were interpolated for US locations, and less than 0.2% of pairs were interpolated for global locations.

For countries or provinces with cities of population larger than 500,000 reported by Demographia, weather and pollution aggregates were produced by performing a weighted average of variables over all cities in the country to avoid data from sparsely populated areas. This affected 137 out of 590 locations from the global dataset (mostly countries). For the remaining global locations, as well as for all US counties, data were drawn from the coordinate of the centroid of that location, which we think is representative of the region given that the vast majority of these locations are sufficiently small and weather variables would not vary significantly within the location.

Population density data was sourced from Demographia (Cox., W, Demographia World Urban Areas, 15^th^ Edition, The Public Purpose), which provided data for urban areas with population greater than 500,000; the United States Census (U.S. Census Bureau, data.census.gov/cedsci); the Iran Statistical Centre; the United Nation’s Projections; City Population (citypopulation.de); and official published estimates for countries not covered by these sources. For data sourced from Demographia, population densities reported are urban densities, whereas other sources primarily reported overall density (spanning urban and non-urban areas). The urban and overall densities are largely on different orders, which weakens the inclusion of population density as an independent variable.

### 2. Estimating the detection delay distribution

Reported data on daily detected COVID-19 infections do not reflect the true infection rate on a given day; rather, it lags behind the true infections due to both the incubation period (during which patients are asymptomatic and less likely to be tested) and the delays between onset of symptoms, testing, and incorporation of test results into official data. We need estimates for the true infection rates for each day to calculate the daily reproduction number (i.e., 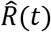), therefore identifying the lag structure between measured infection (*I_M_*) and true infection (*I_N_*), which we call “detection delay” is key to back-tracking from measured infection to estimates of true infection rate.

Prior research has provided several estimates for subsets of overall detection delay. Incubation period, the time between infection to onset of symptoms, has been estimated by several teams.

Li and colleagues (*1*), using data from 10 early patients in China, find the mean incubation period to be 5.2 days, and the delay from onset to first medical visit to be 5.8 days for those infected before January 1^st^ and 4.6 days for the later cases. Lauer and colleagues (*2*) use data from 181 cases to estimate incubation period with mean of 5.5 and median of 5.1, and offer fitted distributions using Lognormal, Gamma, Weibull, and Erlang specifications. In a supplementary graph, they also provide a figure that includes the lags from the onset of symptom to official case detection. Guan et al. (*3*) use data from 291 patients and estimate median incubation period of four days with interquartile range of 2 to 7 days. Linton and colleagues (*4*) use data from 158 cases to estimate the incubation period with a mean (standard deviation) of 5.6 (2.8) days. This delay goes down to 5 (*3*) when Wuhan patients are excluded. They also report onset to hospital admission delay of 3.9 (*3*) days for living patients (155 cases). They provide their full data in an online appendix, where we calculated the onset to case report lag with mean of 5.6 days, median of 5, and standard deviation of 3.8 days. A *New York Times* article (*5*) reports that the Center for Disease Control estimates the lag between onset of symptoms to case detection to be four days. Finally, a Bayesian estimation of the detection delay using abrupt changes in national and state policies by Wibbens and colleagues find the mode of the delay to be 11 days and ranging between 5 and 20 days (*6*).

Overall, these findings are consistent and point to an incubation period of about 5 days and an onset to detection lag of about the same length. We use Lauer et al. estimates for a Lognormal incubation period with parameters 1.62 and 0.418 (leading to mean (standard deviation) of 5.51 (2.4) days), and another Lognormal distribution with parameters 1.47 and 0.52 (resulting in 5 (2.8) days) for onset to detection delay. Combining these two distributions using 10 million Monte-Carlo simulations, we generate the following detection delay lag structure that is used in the analysis. The code calculating this distribution is found at <https://github.com/marichig/weather-conditions-COVID19/>.

**Figure S4:**
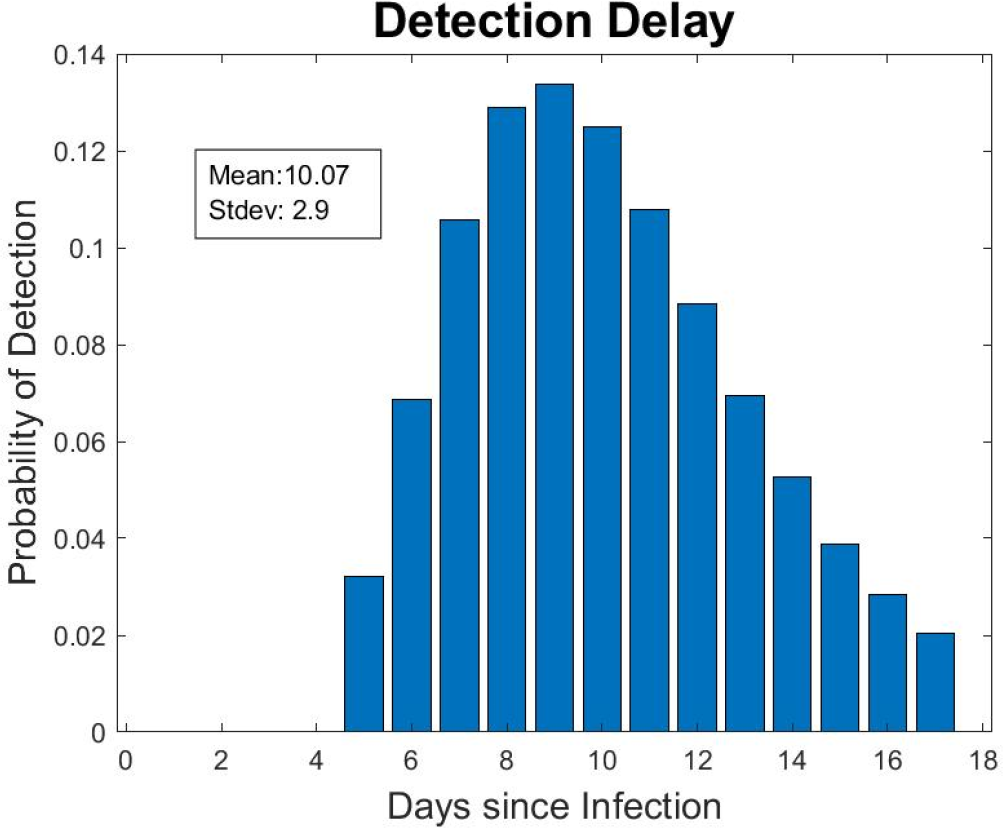
Distribution of Detection Delay.

### 3. Algorithmic estimation of true infection rate

Here we develop an algorithm that provides a more accurate estimation of true exposure than a fixed shift in reported data or averaging data over a time period. We later compare our algorithm’s performance with simpler, more common, methods. We find that accurate estimation of effects of weather variables hinges directly on accurately estimating true infections, making the algorithm in this section key to overall estimation.

Using the delay structure specified in the previous section, one can estimate true infection rates using various methods. The most common solution is to just shift the official infections based on the average, median, or mode of the detection delay (9 to 11 days). This approximation may suffice in steady state but becomes less accurate when estimating time series with exponential growth; the detected infections today are more likely to be from (the many more) recent infections than (the fewer) 10 days ago.

The main objective of our algorithm is to find better estimates for the true infection. This can be seen as deconvolution of detection delay from true infection, when together they produce the observed data. We first calculate the expected number of daily detected cases, given a series of actual infections unknown in the real world. Given the actual infection on day *t, X*(*t*), and the detected infections on day *t, I*(*t*). The following equation would relate the two constructs:

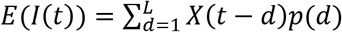

Where *E*(.) takes the expectation on detected infections, *p*(.) is the probability distribution for the detection delay estimated in section 2 of appendix, and the index *d* ranges between 1 to *L* = 17 days to account for different delay lengths. This equation does not account for test coverage, but as discussed below test coverage cancels out of the final reproduction number calculations, and as such only impacts variability of outcomes, otherwise having limited impact on results. Note that this equation is under-specified; for one value of the known measure *I*, one has to find up to *L* values of the unknown *X* (in our case, no detection is expected in the first 4 days, so L-4=13 values of *X* contribute to a value of *I* (Figure S4)). However, given the overlap on *X*’s determining subsequent *I* values, the system of equations connecting *I* and *X* values for *I* time series extending over *T* days would include *T* known values (for *I*) and *T + L* unknown *X* values. Different approaches could then be pursued to find approximate solutions for this system of equations.

Using exact Maximum Likelihood suffers from intractability of specifying the Likelihood for highly correlated Poisson distributions (Poisson is a natural alternative in this case). We compared two alternatives, one using Normally distributed approximations for *I* as a function of *X*, and another using a direct minimization of the gap between *I* values and their expectation. The latter proves both simpler conceptually and more accurate in synthetic data, so we picked that for the main analysis:

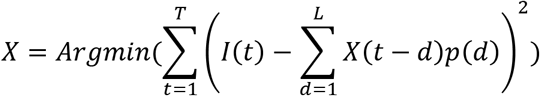

Given the underspecification of original system of equations, this optimization will include many solutions. To identify a more realistic solution from that set, we add a regularization term that penalizes the gap between subsequent values for *X*. Specifically, we use the following optimization:

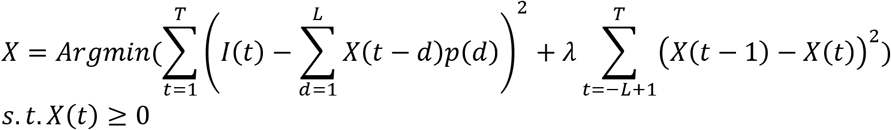

The solution to this optimization can be found using standard quadratic programming methods, allowing for fast and scalable solutions. We conducted sensitivity analysis to find the regularization parameter, λ, offering the best overall ability of the algorithm to find true infections in synthetic data. The algorithm that works well is with λ values in the 0.1 to 0.5 range and not very sensitive to exact value; we used a value of 0.2 in our analysis. An implementation of this code in MATLAB is available from <https://github.com/marichig/weather-conditions-COVID19/>.

For each location in our dataset, we used this algorithm to estimate the true infections (Î*_N_*(*t*) *= X*(*t*)), on a daily basis, starting from 17 days before the first detected infection, and stopping 5 days before the last day with data (because only infections from 5 days or further back could be found in current measures of infection; see the detection delay distribution (Figure S4)). These values were then used to create the dependent variable, 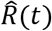, as discussed in the body of the article:

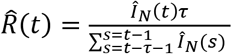

We recognize that not all infections are reported, and a large fraction may remain unknown. Assuming that only a fraction *f* (0 ≤ *f* ≤ 1) of actual infections are reported, *I_M_* would be *f* of total infections that could have been detected on a given day, and estimated *Î_N_*(*t*) will be the fraction *f* of true infections as a result. While these underestimations are likely very significant if we cared about absolute values of *Î_N_*(*t*), note that *Î_N_*(*t*) values show up both in the numerator and denominator of 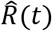 equation. Therefore, multiplying both by a fixed constant makes no difference in the estimated 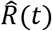.

We also recognize that, early on during the infection, *f* may increase with expanding test capacity until reaching a steady state value. Therefore, as later discussed, we drop the first few data points for each region and check the sensitivity of the result to dropping fewer or more days. Finally, our synthetic analysis (section 5.2.2.3, Experiment 10) shows results are robust to various trajectories for *f* over the course of epidemic.

### 4. Statistical sensitivity analyses and robustness checks

In our main specification we included weather predictors that are (a) of theoretical interest and (b) do not cause a collinearity issue when included together (correlations among main weather variables are reported in Table S2). Here we report eight different sets of sensitivity tests that assess the robustness of our findings to various assumptions, boundary conditions and inclusion of other weather variables.^1^ Here is a summary of the results, before we go into the details: (*1*) In our main specification we used a delay of τ=20 days from exposure to virus to resolution (recovery or death) to calculate reproduction number R_0_. Here we tested the robustness of our results over a spectrum of reasonable durations of delay from 15 to 25 days, finding no major impact on the results. (*2*) We tested our main specification under five additional exclusion criteria and specifications: exclusion of the last few days of data (for which true infection estimates may be less reliable), exclusions of top 1% R_0_ of our sample (which may be generated due to reporting issues), exclusions of non-US sample, inclusion of date fixed effects (to control for the possible global events impacting outcomes) and inclusion of location-specific quadratic trend effects (to control for the possible non-linearity in each location’s response over time). The key results on mean temperature, ultraviolet index and precipitation are robust across all specifications. (*3*) In our main specification we excluded the first 20 days since new infection exceeds 1 for each location to account for early-on changes in test coverage and to get stable estimates for reproduction number. Here we tested the robustness of our results to other exclusion periods, ranging from first 10 days to 30 days, finding no major impact on the key results. (*4*) To exclude the possibility that our results are driven by mechanical features of our variable construction and model specification, we used a set of placebo weather variables, which are randomly permuted across locations and shifted over a specific number of days, and re-estimated our main models using these placebo weather variables. We found few significant effects under these placebo tests. (*5*) In our main specification we chose a linear spline effect of mean temperature with a knot at 25 degrees. Here we tested how our results are sensitive to different choices of knots, finding the 25 degree provides the best fit. (*6*) In our main specification we found a somewhat counterintuitive U-shaped effect of ultraviolet index. To ensure the effect is not driven by observations with extremely large UV indices, we excluded observations with top 5% and 10% UV index values in our sample and repeated our analysis. The U-shaped effect of UV is robust to these exclusions. (*7*) We report analyses that include several interaction terms, as well as some additional weather variables of interest (e.g., absolute humidity, NO_2_ and PM_2.5_). Our main results are robust to these inclusions and we did not find significant effects of these additional weather variables. (*8*) Finally, we assess the overall robustness of projections of relative predicted risk (CRW) using a series of different specifications, from using US-only samples, to subsets of specifications reported in other analyses. Projections using these different measures are highly consistent across these specifications (with correlations above 0.9 in most cases).

**Table S2.** Correlations among weather variables in the main specification

| | $\text{SO}_2$ (log) | $\text{O}_3$ (log) | Windspeed (log) | Pressure | Precipitation (log) | Relative Humidity | Mean temperature | UV index | Diurnal temperature |
| --- | --- | --- | --- | --- | --- | --- | --- | --- | --- |
| Sulfur dioxide (log) | 1.00 |  |  |  |  |  |  |  |  |
| Ozone (log) | -0.51 | 1.00 |  |  |  |  |  |  |  |
| Windspeed (log) | -0.23 | 0.27 | 1.00 |  |  |  |  |  |  |
| Pressure | 0.19 | -0.14 | -0.29 | 1.00 |  |  |  |  |  |
| Precipitation (log) | -0.14 | 0.08 | 0.16 | -0.36 | 1.00 |  |  |  |  |
| Relative Humidity | -0.16 | -0.07 | -0.01 | -0.18 | 0.53 | 1.00 |  |  |  |
| Mean temperature | 0.13 | -0.07 | -0.06 | -0.31 | 0.01 | -0.05 | 1.00 |  |  |
| UV index | 0.04 | 0.00 | -0.05 | -0.22 | 0.08 | 0.01 | 0.79 | 1.00 |  |
| Diurnal temperature | -0.02 | -0.03 | -0.29 | 0.10 | -0.28 | -0.27 | 0.07 | 0.10 | 1.00 |

#### 4.1. Sensitivity to duration of disease (15, 20, 25)

Table S3 presents the results using our main specification with R_0_ calculated from 15, 20 and 25 days of delay respectively. The coefficients and significance level for each weather variable are largely unchanged and consistent across different durations, especially for ozone, precipitation, mean temperature and ultraviolet index.

**Table S3.**
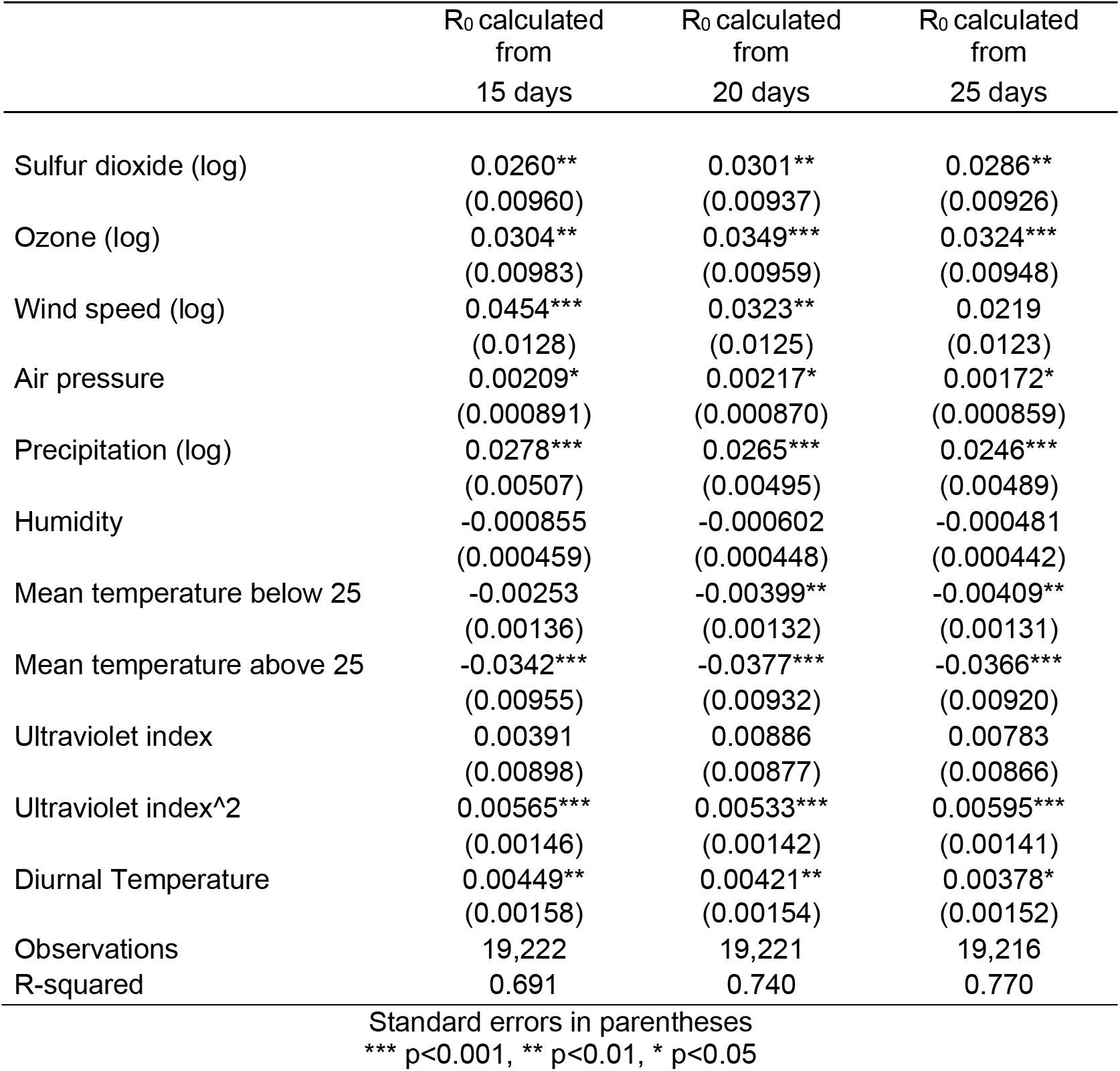
Regression results with various duration of delay to calculate R_0_

| | $R_0$ calculated<br>from<br>15 days | $R_0$ calculated<br>from<br>20 days | $R_0$ calculated<br>from<br>25 days |
| --- | --- | --- | --- |
| Sulfur dioxide (log) | 0.0260**<br>(0.00960) | 0.0301**<br>(0.00937) | 0.0286**<br>(0.00926) |
| Ozone (log) | 0.0304**<br>(0.00983) | 0.0349***<br>(0.00959) | 0.0324***<br>(0.00948) |
| Wind speed (log) | 0.0454***<br>(0.0128) | 0.0323**<br>(0.0125) | 0.0219<br>(0.0123) |
| Air pressure | 0.00209*<br>(0.000891) | 0.00217*<br>(0.000870) | 0.00172*<br>(0.000859) |
| Precipitation (log) | 0.0278***<br>(0.00507) | 0.0265***<br>(0.00495) | 0.0246***<br>(0.00489) |
| Humidity | -0.000855<br>(0.000459) | -0.000602<br>(0.000448) | -0.000481<br>(0.000442) |
| Mean temperature below 25 | -0.00253<br>(0.00136) | -0.00399**<br>(0.00132) | -0.00409**<br>(0.00131) |
| Mean temperature above 25 | -0.0342***<br>(0.00955) | -0.0377***<br>(0.00932) | -0.0366***<br>(0.00920) |
| Ultraviolet index | 0.00391<br>(0.00898) | 0.00886<br>(0.00877) | 0.00783<br>(0.00866) |
| Ultraviolet index^2 | 0.00565***<br>(0.00146) | 0.00533***<br>(0.00142) | 0.00595***<br>(0.00141) |
| Diurnal Temperature | 0.00449**<br>(0.00158) | 0.00421**<br>(0.00154) | 0.00378*<br>(0.00152) |
| Observations | 19,222 | 19,221 | 19,216 |
| R-squared | 0.691 | 0.740 | 0.770 |

Standard errors in parentheses
\*\*\* $p < 0.001$ , \*\* $p < 0.01$ , \* $p < 0.05$

#### 4.2. Sensitivity to exclusion criteria and including additional controls

Table S4 presents the results when we excluded the last 4 days of our data (19% of the total sample in our main specification), top 1% R_0_, used a US-only sample, or included date fixed effects or location-specific quadratic trend effects. Across specifications we observed robust and consistent estimates of the positive effect of precipitation, linear spline effect of mean temperature, and U-shaped effect of ultraviolet index. Other weather effects are somewhat less robust; the positive effects of air pollutants (i.e., SO_2_ and O_3_), while robust to excluding last 4 days of data, extreme R_0_, and the inclusion of date fixed effects, went away when we only used US locations or with the inclusion of location-specific quadratic trends. The positive effect of wind speed, while robust to both exclusion criteria, is no longer significant with additional controls. The positive effects of air pressure and diurnal temperature are only robust to using US data and excluding extreme R_0_, but not to other tests. We note that including a quadratic trend term adds another location-specific parameter, which would further absorb variations in weather and air-pollutants in each location, and as such is expected to attenuate the parameter estimates further.

**Table S4.** Regression results with various exclusion criteria and specifications

| | Exclude last 4<br>days of data | Exclude top 1%<br>$R_0$ | Only using US<br>sample | Including date<br>fixed effects | Including<br>location-specific<br>quadratic trend |
| --- | --- | --- | --- | --- | --- |
| Sulfur dioxide (log) | 0.0276**<br>(0.00998) | 0.0287**<br>(0.00928) | 0.00756<br>(0.0112) | 0.0302**<br>(0.00931) | 0.0149<br>(0.00804) |
| Ozone (log) | 0.0234*<br>(0.00991) | 0.0367***<br>(0.00953) | 0.0163<br>(0.0112) | 0.0287**<br>(0.00948) | 0.0148<br>(0.00822) |
| Wind speed (log) | 0.0422**<br>(0.0132) | 0.0348**<br>(0.0124) | 0.0455**<br>(0.0146) | 0.00555<br>(0.0126) | 0.00541<br>(0.0107) |
| Air pressure | 0.00175<br>(0.000948) | 0.00261**<br>(0.000867) | 0.00308**<br>(0.00103) | -0.00138<br>(0.000905) | -0.00109<br>(0.000799) |
| Precipitation (log) | 0.0322***<br>(0.00531) | 0.0258***<br>(0.00491) | 0.0312***<br>(0.00531) | 0.0155**<br>(0.00496) | 0.0195***<br>(0.00428) |
| Humidity | -0.000836<br>(0.000489) | -0.000464<br>(0.000444) | 6.02e-05<br>(0.000488) | -0.000221<br>(0.000459) | -0.000814*<br>(0.000395) |
| Mean temperature below 25 | -0.000286<br>(0.00144) | -0.00406**<br>(0.00131) | -0.00533***<br>(0.00143) | -0.00577***<br>(0.00145) | -0.00269*<br>(0.00117) |
| Mean temperature above 25 | -0.0374***<br>(0.00999) | -0.0371***<br>(0.00928) | -0.0555***<br>(0.0134) | -0.0384***<br>(0.00913) | -0.0275***<br>(0.00805) |
| Ultraviolet index | -0.00523<br>(0.00988) | 0.00952<br>(0.00868) | -0.000405<br>(0.00985) | 0.00199<br>(0.00904) | 0.00288<br>(0.00772) |
| Ultraviolet index^2 | 0.00519***<br>(0.00155) | 0.00528***<br>(0.00142) | 0.00882***<br>(0.00233) | 0.00467**<br>(0.00142) | 0.00540***<br>(0.00128) |
| Diurnal Temperature | 0.00302<br>(0.00168) | 0.00425**<br>(0.00153) | 0.00553***<br>(0.00160) | 0.00304<br>(0.00157) | 0.000710<br>(0.00132) |
| Observations | 15,595 | 19,031 | 13,953 | 19,216 | 19,221 |
| R-squared | 0.771 | 0.730 | 0.720 | 0.778 | 0.840 |

Standard errors in parentheses
\*\*\* $p < 0.001$ , \*\* $p < 0.01$ , \* $p < 0.05$

#### 4.3. Sensitivity to shifting the data start date after first exposure

Table S5 presents the results when we excluded first 10, 15, 20, 25, and 30 days since new infection exceeds 1 for the first time in each location. Overall, we observed robust and consistent estimates of the positive effect of precipitation, U-shaped effect of UV index, and linear spline effect of mean temperature, except when we excluded the first 30 days of data for each location, where we would lose more than half of our sample as compared to the main specification (first 20 days excluded). It is possible that by constraining our estimation on later periods, we are focusing on periods when lockdown and social distancing are fully in effect and thus there are few variations left in R_0_ that can be explained by environmental factors. However, the coefficients for these effects are still consistent and have the same sign. For example, with the first 30 days excluded, we estimated that with a one degree increase in mean temperature after 25 degrees, the estimated R_0_ will still decrease by ~2%. The positive effects of SO_2_, O_3_, wind speed, air pressure, and diurnal temperature were less robust and no longer significant when we excluded the first 25 or 30 days.

**Table S5.** Regression results with various exclusion criteria for initial periods

|  | Exclude first 10<br>days | Exclude first 15<br>days | Exclude first 20<br>days | Exclude first 25<br>days | Exclude first 30<br>days |
| --- | --- | --- | --- | --- | --- |
| Sulfur dioxide (log) | 0.00558<br>(0.00777) | 0.0185*<br>(0.00858) | 0.0301**<br>(0.00937) | 0.0220*<br>(0.0106) | 0.0169<br>(0.0128) |
| Ozone (log) | 0.00977<br>(0.00780) | 0.0192*<br>(0.00872) | 0.0349***<br>(0.00959) | 0.0317**<br>(0.0110) | 0.0217<br>(0.0129) |
| Wind speed (log) | 0.0935***<br>(0.0100) | 0.0853***<br>(0.0112) | 0.0323**<br>(0.0125) | 0.00949<br>(0.0143) | -0.00411<br>(0.0175) |
| Air pressure | 0.00853***<br>(0.000696) | 0.00652***<br>(0.000786) | 0.00217*<br>(0.000870) | 0.000893<br>(0.000989) | 0.00164<br>(0.00119) |
| Precipitation (log) | 0.0325***<br>(0.00390) | 0.0345***<br>(0.00441) | 0.0265***<br>(0.00495) | 0.0181**<br>(0.00572) | 0.0216**<br>(0.00704) |
| Humidity | -0.000526<br>(0.000360) | -0.000740<br>(0.000403) | -0.000602<br>(0.000448) | -9.62e-05<br>(0.000515) | -0.000353<br>(0.000637) |
| Mean temperature below 25 | -0.000714<br>(0.00108) | -0.00316**<br>(0.00120) | -0.00399**<br>(0.00132) | -0.00434**<br>(0.00153) | -0.00215<br>(0.00192) |
| Mean temperature above 25 | -0.0208*<br>(0.00873) | -0.0299**<br>(0.00919) | -0.0377***<br>(0.00932) | -0.0370***<br>(0.00990) | -0.0190<br>(0.0110) |
| Ultraviolet index | 0.0135<br>(0.00712) | 0.00740<br>(0.00793) | 0.00886<br>(0.00877) | 0.0125<br>(0.00990) | 0.00677<br>(0.0119) |
| Ultraviolet index^2 | 0.00731***<br>(0.00121) | 0.00789***<br>(0.00132) | 0.00533***<br>(0.00142) | 0.00639***<br>(0.00158) | 0.00826***<br>(0.00183) |
| Diurnal Temperature | 0.00661***<br>(0.00123) | 0.00707***<br>(0.00138) | 0.00421**<br>(0.00154) | 0.00389*<br>(0.00177) | 0.00241<br>(0.00219) |
| Observations | 32,232 | 25,913 | 19,221 | 13,670 | 9,071 |
| R-squared | 0.699 | 0.705 | 0.740 | 0.779 | 0.814 |

Standard errors in parentheses
\*\*\* p<0.001, \*\* p<0.01, \* p<0.05

#### 4.4. Placebo tests (random shifts of weather data)

Placebo tests allow us to ensure that mechanical features of the statistical estimation method are not driving any of the results. The basic intuition is simple: if we feed to the algorithm independent variables that are not matched correctly to the estimated exposure rates, we should not observe any major correlations. To implement, we first randomly permuted weather variables across locations in our data, and then shifted all weather variables in each location to earlier periods by a specific number of days, where the number is randomly drawn from a uniform distribution U(0,300). We then performed the statistical analysis using these placebo weather variables. As shown in Table S6, most of the weather effects are completely gone, especially in our main specification where first 20 days are excluded. The only exception is when we observe a negative and significant linear effect of ultraviolet index when first 10 days are excluded. A single “significant” effect at p=0.05 out of over 50 estimated coefficients is expected based on chance alone.

We also note the relatively large R-squared values in these placebo tests: fixed effects and location trends provide much explanatory power regardless of weather and pollution. This observation informed our choice to focus our statistical models on simpler and more interpretable linear forms rather than using cross-validation or other prediction-driven methods to specify terms and functional forms for statistical models.

**Table S6.**
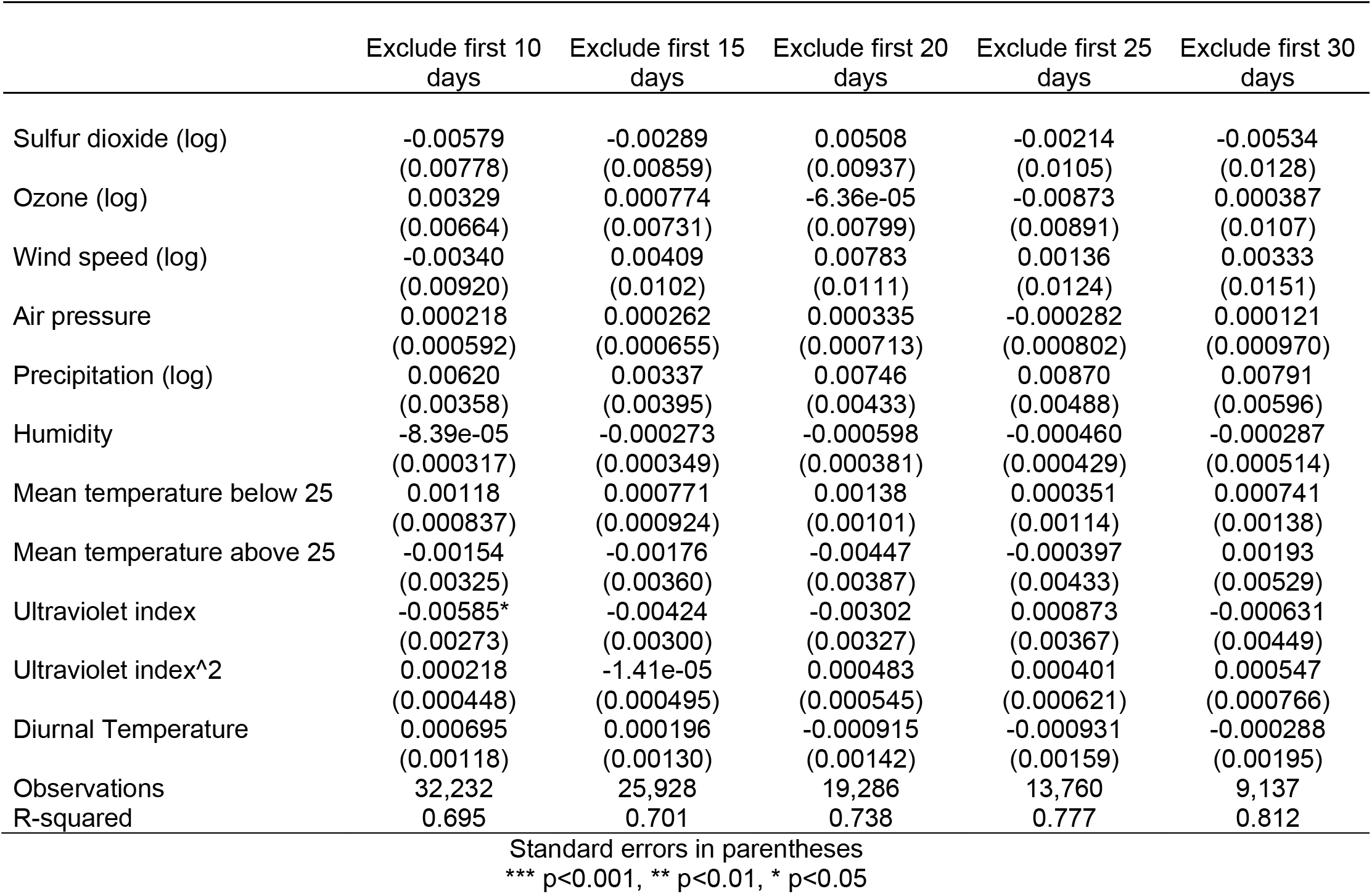
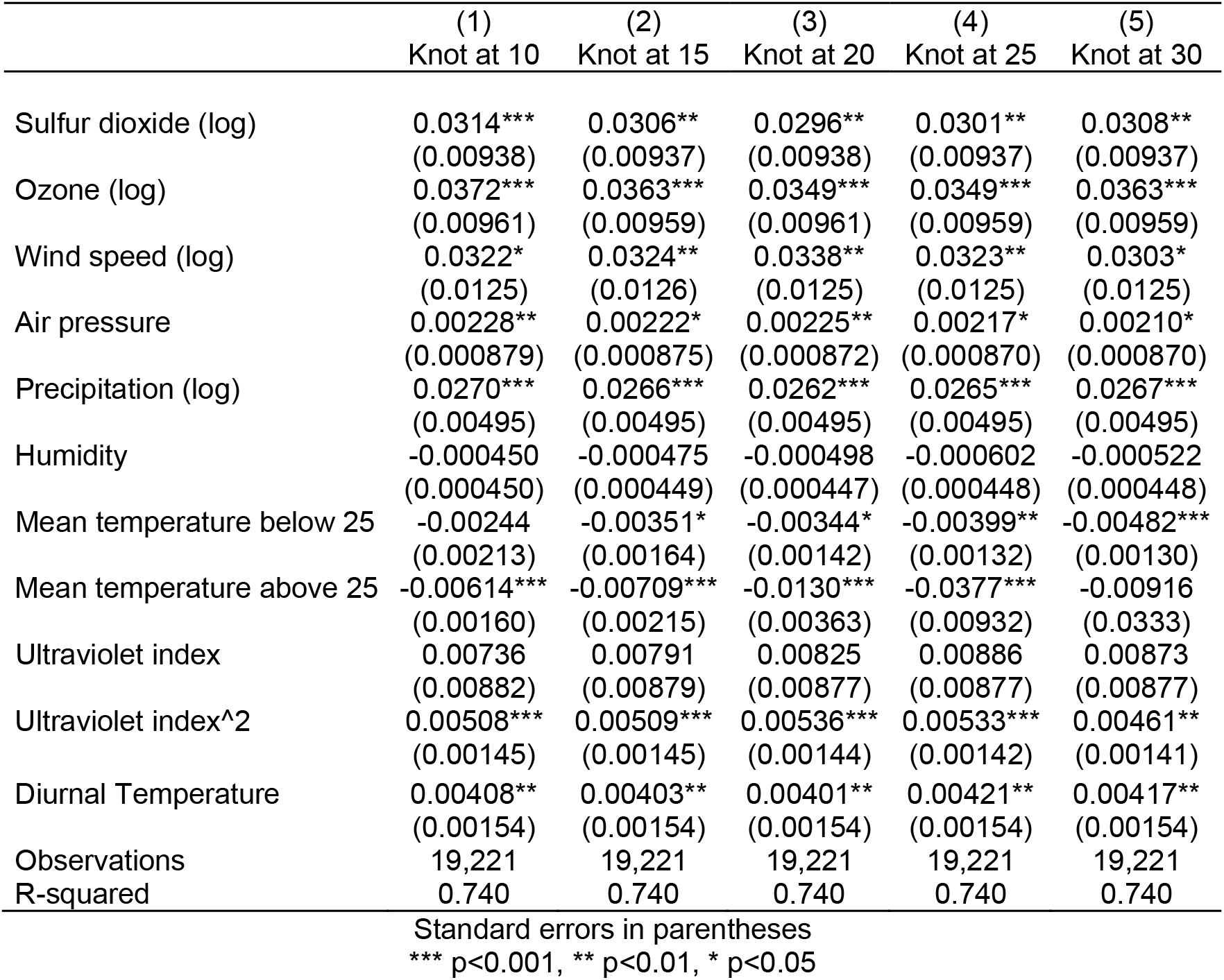
Regression results using placebo weather with various exclusion criteria for initial periods

#### 4.5. Different knot for linear spline effect of mean temperature

In the main specification we used a linear spline effect of mean temperature with a knot at 25 degrees as it provides better fit than linear or quadratic effect of temperature. Here we test the sensitivity of our results to the choice of knots over a wide range of mean temperatures from −15 degrees to 30 degrees. As shown in Table S7, the temperature effect after the knot is statistically significant and much larger at 25 degrees (−.0377, p<.001) than knots at other degrees. Hence, we chose the knot at 25 degrees as our main specification. Given the limited number of locations with temperature above 30 in our estimation sample, increasing the knot beyond that level is not supported by our data.

**Table S7.** Regression results with different knots for linear spline effect of mean temperature

|  | (1)<br>Knot at -15 | (2)<br>Knot at -10 | (3)<br>Knot at -5 | (4)<br>Knot at 0 | (5)<br>Knot at 5 |
| --- | --- | --- | --- | --- | --- |
| Sulfur dioxide (log) | 0.0308**<br>(0.00937) | 0.0306**<br>(0.00937) | 0.0309***<br>(0.00937) | 0.0311***<br>(0.00937) | 0.0312***<br>(0.00939) |
| Ozone (log) | 0.0362***<br>(0.00959) | 0.0360***<br>(0.00959) | 0.0363***<br>(0.00959) | 0.0366***<br>(0.00959) | 0.0367***<br>(0.00960) |
| Wind speed (log) | 0.0302*<br>(0.0125) | 0.0300*<br>(0.0125) | 0.0298*<br>(0.0125) | 0.0302*<br>(0.0125) | 0.0307*<br>(0.0125) |
| Air pressure | 0.00206*<br>(0.000870) | 0.00202*<br>(0.000870) | 0.00205*<br>(0.000870) | 0.00210*<br>(0.000870) | 0.00216*<br>(0.000875) |
| Precipitation (log) | 0.0266***<br>(0.00495) | 0.0266***<br>(0.00495) | 0.0268***<br>(0.00495) | 0.0269***<br>(0.00495) | 0.0269***<br>(0.00496) |
| Humidity | -0.000525<br>(0.000447) | -0.000515<br>(0.000447) | -0.000491<br>(0.000447) | -0.000470<br>(0.000449) | -0.000486<br>(0.000450) |
| Mean temperature below 25 | 0.166*<br>(0.0823) | 0.0518**<br>(0.0186) | 0.0177*<br>(0.00873) | 0.00194<br>(0.00526) | -0.00282<br>(0.00325) |
| Mean temperature above 25 | -0.00498***<br>(0.00131) | -0.00528***<br>(0.00131) | -0.00541***<br>(0.00132) | -0.00525***<br>(0.00134) | -0.00517***<br>(0.00140) |
| Ultraviolet index | 0.00909<br>(0.00877) | 0.00958<br>(0.00877) | 0.00950<br>(0.00877) | 0.00888<br>(0.00877) | 0.00850<br>(0.00878) |
| Ultraviolet index^2 | 0.00462**<br>(0.00141) | 0.00464***<br>(0.00141) | 0.00464***<br>(0.00141) | 0.00466***<br>(0.00141) | 0.00472***<br>(0.00142) |
| Diurnal Temperature | 0.00418**<br>(0.00154) | 0.00434**<br>(0.00154) | 0.00438**<br>(0.00154) | 0.00430**<br>(0.00154) | 0.00420**<br>(0.00154) |
| Observations | 19,221 | 19,221 | 19,221 | 19,221 | 19,221 |
| R-squared | 0.740 | 0.740 | 0.740 | 0.740 | 0.740 |

#### 4.6 Excluding observations with high ultraviolet index

Table S8 and Table S9 report the estimates of UV index effects when excluding observations in the top 5% (13.25) and top 10% (10.66) UV index in our sample. Results show that the U-shaped effect of UV index is significant and consistent across specifications and largely robust to the exclusion of observations with large UV index, showing that our results are not solely driven by extreme values. In fact, if anything, the curvature of UV impact becomes sharper (more positive) when we exclude those extreme values. While we lack a theoretically strong justification for the increasing part of the U-shaped effect, one could speculate that it relates to shift of social interactions to riskier indoor environments when UV index is very high. We cannot directly test this or other alternative hypotheses in our data.

**Table S8.** Regression results excluding observations with top 5% UV index under various exclusion criteria for initial periods

|  | Exclude first 10<br>days | Exclude first 15<br>days | Exclude first 20<br>days | Exclude first 25<br>days | Exclude<br>first 30<br>days |
| --- | --- | --- | --- | --- | --- |
| Sulfur dioxide (log) | 0.00194<br>(0.00788) | 0.0144<br>(0.00874) | 0.0249**<br>(0.00962) | 0.0180<br>(0.0110) | 0.0117<br>(0.0133) |
| Ozone (log) | 0.00564<br>(0.00791) | 0.0147<br>(0.00888) | 0.0323**<br>(0.00985) | 0.0310**<br>(0.0114) | 0.0202<br>(0.0135) |
| Wind speed (log) | 0.0950***<br>(0.0100) | 0.0879***<br>(0.0112) | 0.0339**<br>(0.0125) | 0.0116<br>(0.0145) | 0.000673<br>(0.0177) |
| Air pressure | 0.00857***<br>(0.000692) | 0.00665***<br>(0.000782) | 0.00228**<br>(0.000868) | 0.000923<br>(0.000990) | 0.00177<br>(0.00119) |
| Precipitation (log) | 0.0351***<br>(0.00393) | 0.0372***<br>(0.00446) | 0.0281***<br>(0.00503) | 0.0191**<br>(0.00584) | 0.0229**<br>(0.00725) |
| Humidity | -0.000653<br>(0.000360) | -0.000850*<br>(0.000403) | -0.000701<br>(0.000450) | -0.000116<br>(0.000519) | -0.000441<br>(0.000643) |
| Mean temperature<br>below 25 | -0.000564<br>(0.00108) | -0.00285*<br>(0.00119) | -0.00364**<br>(0.00132) | -0.00406**<br>(0.00152) | -0.00186<br>(0.00192) |
| Mean temperature<br>above 25 | -0.0455***<br>(0.0100) | -0.0479***<br>(0.0104) | -0.0478***<br>(0.0104) | -0.0439***<br>(0.0109) | -0.0194<br>(0.0120) |
| Ultraviolet index | 0.0196**<br>(0.00734) | 0.00976<br>(0.00814) | 0.00809<br>(0.00901) | 0.00777<br>(0.0102) | 0.00290<br>(0.0124) |
| Ultraviolet index^2 | 0.0117***<br>(0.00163) | 0.0106***<br>(0.00180) | 0.00594**<br>(0.00199) | 0.00466*<br>(0.00223) | 0.00774**<br>(0.00270) |
| Diurnal<br>Temperature | 0.00714***<br>(0.00123) | 0.00753***<br>(0.00138) | 0.00451**<br>(0.00154) | 0.00426*<br>(0.00178) | 0.00334<br>(0.00221) |
| Observations | 30,871 | 24,738 | 18,259 | 12,915 | 8,477 |
| R-squared | 0.706 | 0.711 | 0.743 | 0.780 | 0.816 |

Standard errors in parentheses
\*\*\* p<0.001, \*\* p<0.01, \* p<0.05

**Table S9.** Regression results excluding observations with top 10% UV index under various exclusion criteria for initial periods

|  | Exclude first<br>10 days | Exclude first<br>15 days | Exclude first<br>20 days | Exclude first<br>25 days | Exclude first<br>30 days |
| --- | --- | --- | --- | --- | --- |
| Sulfur dioxide (log) | 0.0243*<br>(0.00994) | 0.0243*<br>(0.00994) | 0.0243*<br>(0.00994) | 0.0144<br>(0.0114) | 0.0122<br>(0.0140) |
| Ozone (log) | 0.0319**<br>(0.0101) | 0.0319**<br>(0.0101) | 0.0319**<br>(0.0101) | 0.0289*<br>(0.0118) | 0.0188<br>(0.0141) |
| Wind speed (log) | 0.0309*<br>(0.0128) | 0.0309*<br>(0.0128) | 0.0309*<br>(0.0128) | 0.0120<br>(0.0147) | -0.00624<br>(0.0180) |
| Air pressure | 0.00256**<br>(0.000877) | 0.00256**<br>(0.000877) | 0.00256**<br>(0.000877) | 0.00140<br>(0.000994) | 0.00199<br>(0.00120) |
| Precipitation (log) | 0.0309***<br>(0.00516) | 0.0309***<br>(0.00516) | 0.0309***<br>(0.00516) | 0.0229***<br>(0.00599) | 0.0284***<br>(0.00749) |
| Humidity | -0.000810<br>(0.000457) | -0.000810<br>(0.000457) | -0.000810<br>(0.000457) | -0.000296<br>(0.000526) | -0.000769<br>(0.000655) |
| Mean temperature<br>below 25 | -0.00335*<br>(0.00133) | -0.00335*<br>(0.00133) | -0.00335*<br>(0.00133) | -0.00370*<br>(0.00153) | -0.00179<br>(0.00193) |
| Mean temperature<br>above 25 | -0.0657***<br>(0.0168) | -0.0657***<br>(0.0168) | -0.0657***<br>(0.0168) | -0.0507**<br>(0.0178) | -0.0414*<br>(0.0204) |
| Ultraviolet index | 0.0104<br>(0.00948) | 0.0104<br>(0.00948) | 0.0104<br>(0.00948) | 0.00857<br>(0.0108) | 0.0104<br>(0.0132) |
| Ultraviolet index^2 | 0.00709**<br>(0.00231) | 0.00709**<br>(0.00231) | 0.00709**<br>(0.00231) | 0.00531*<br>(0.00261) | 0.0106***<br>(0.00320) |
| Diurnal<br>Temperature | 0.00477**<br>(0.00157) | 0.00477**<br>(0.00157) | 0.00477**<br>(0.00157) | 0.00477**<br>(0.00180) | 0.00371<br>(0.00225) |
| Observations | 17,290 | 17,290 | 17,290 | 12,115 | 7,843 |
| R-squared | 0.740 | 0.740 | 0.740 | 0.779 | 0.813 |

Standard errors in parentheses
\*\*\* p<0.001, \*\* p<0.01, \* p<0.05

#### 4.7 Inclusion of interaction terms and additional variables

Table S10 presents the results of including additional weather effects and interaction terms in our main specification. We first included additional weather effects of absolute humidity, nitrogen dioxide, particulate matter, visibility, wind direction, cloud cover, snow, and moon illumination. We did not observe significant effects of these additional weather variables, except for moon illumination, where we observed a significant negative effect. But most of our main results are robust to the inclusion of additional variables, except for wind speed and air pressure, which are no longer significant with the inclusion of moon illumination. We then explored some interaction effects between weather variables. We found a significant negative interaction between the quadratic term of UV index and precipitation, indicating the U-shaped effect of UV index will be dampened with the increase of precipitation. We also found a positive and significant interaction between mean temperature above 25°C and sulfur dioxide, indicating the negative effect of temperature will be attenuated by increased sulfur dioxide level. Finally, we observed a negative and significant interaction between particulate matter and air pressure, indicating the positive effect of particulate matter will attenuate with higher air pressure. While we do not have good theoretical explanations for the moon illumination effect and these interaction effects, we reported them here to show other weather effects are robust to this additional inclusion and point to possible avenues for further investigation.

**Table S10.**
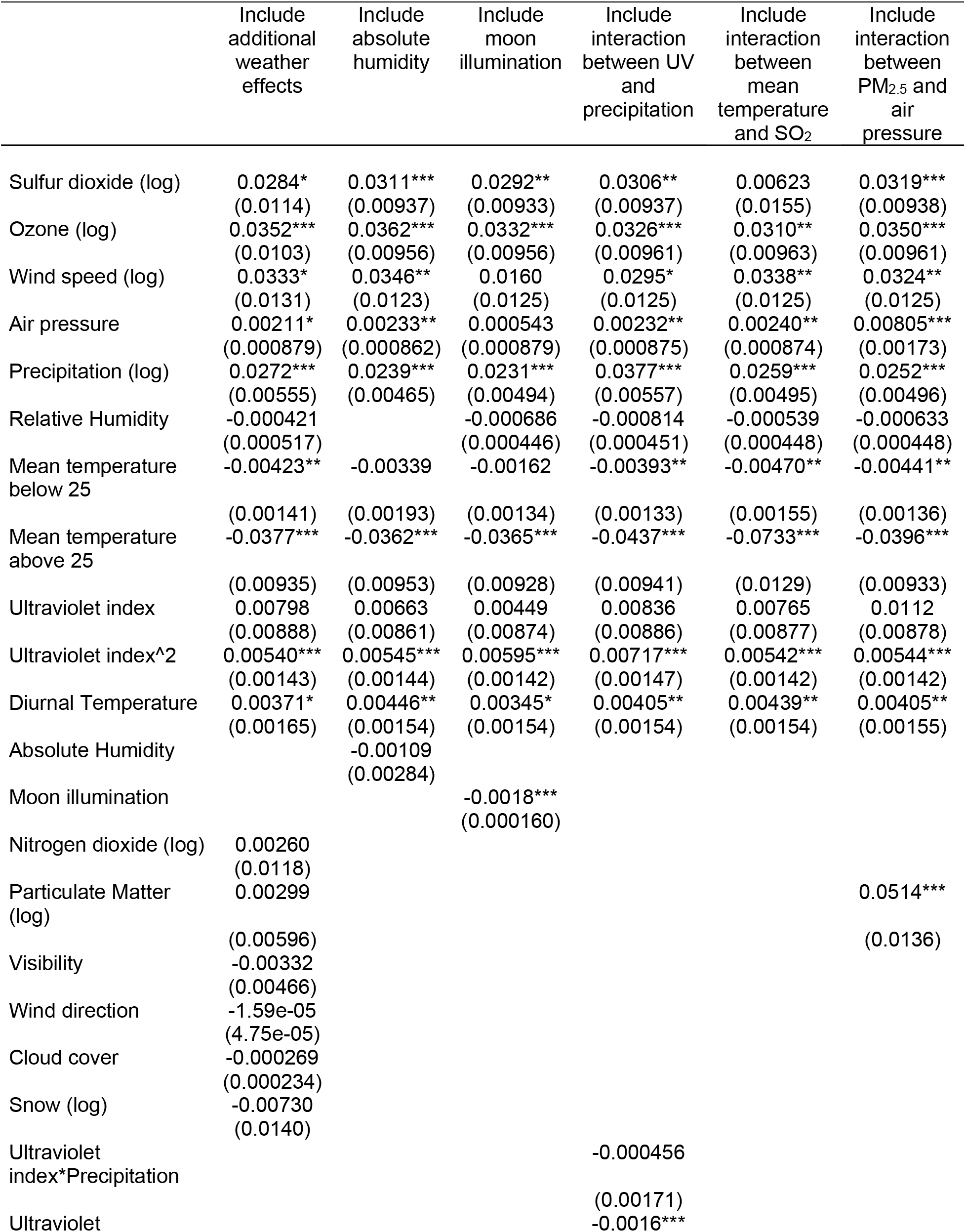
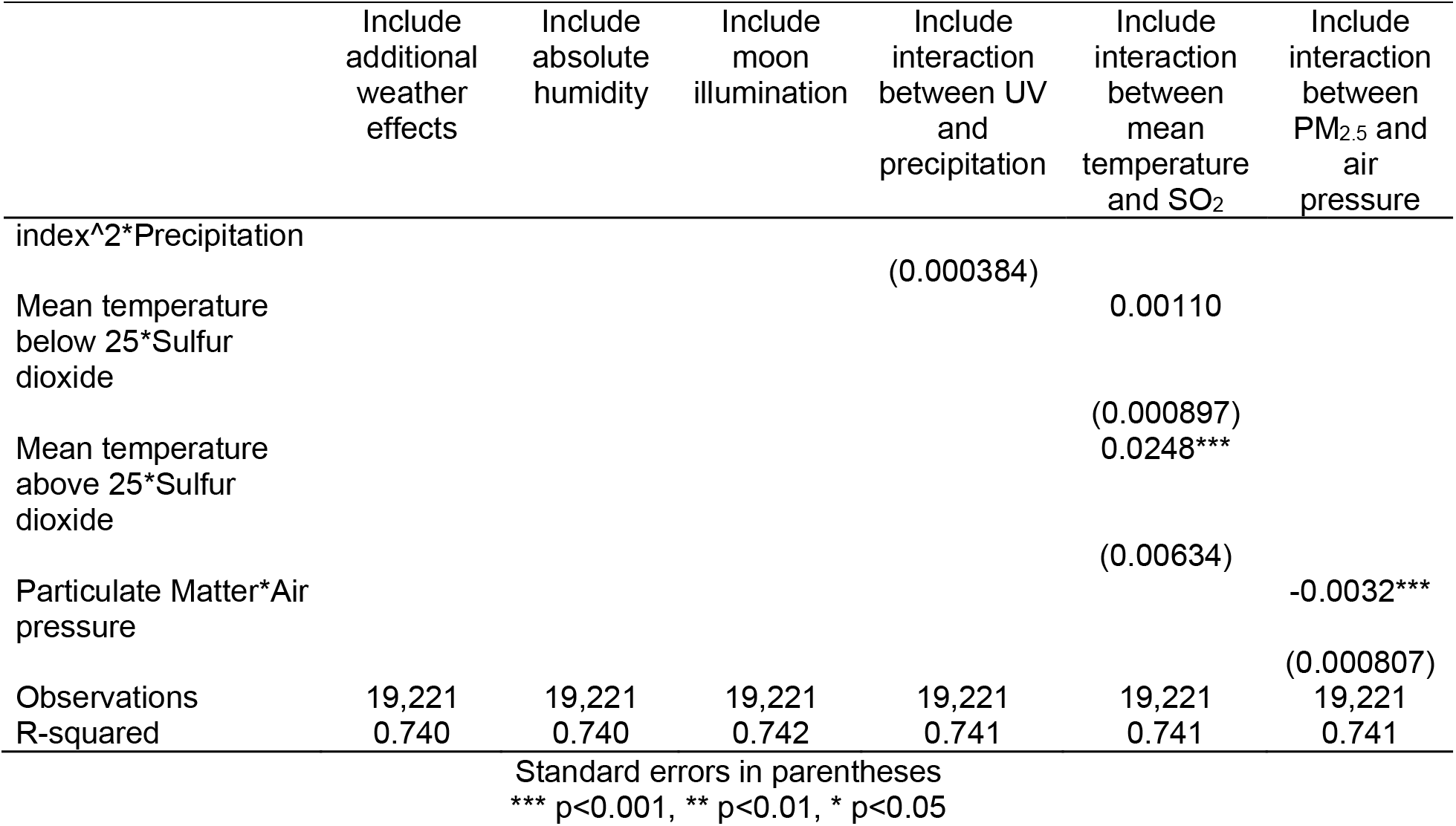
Regression results including additional interaction terms and weather effects

#### 4.8 Overall robustness of projections to various specifications

Our last test focuses on the overall robustness of projections to alternative specifications. To do this we constructed a sample independent from estimation data consisting of 1072 cities and calculated their relative predicted risk (based on weather and air pollution vector from January 23 2019 to January 23, 2020) relative to the median of predicted risk in our estimation sample across 9 alternative specifications. These predictions are similar to CRW measures we report with two notable caveats: first, we use daily, rather than 15-day, averages for this set of projections. The use of much more variable daily inputs will significantly increase the variance in predictions and elicit any differences between alternative models more clearly. Second, we use the median, rather than 95^th^ percentile of the estimation sample, to normalize these measures so that the comparisons are centered around the same point at the value of 1.

We then calculate correlations and mean absolute errors (MAE) between projections from several alternative models and report them in Table S11 (see the model specifications compared below the table). Results show that there are high correlations (average correlation =.945, SD=.034) and low MAEs (average MAE=.047, SD=.023) across various CRWs, showing that risk projections and our results are robust to different exclusion criteria and inclusion of additional variables.

**Table S11.** Correlation and mean absolute error of relative risk projection for 1072 urban cities over 1 year across 9 specifications (correlation reported in lower diagonal (blue), MAE between specifications in upper diagonal (green), MAE against a constant value of 1 on the diagonal (white))

|  | A | B | C | D | E | F | G | H | I |
| --- | --- | --- | --- | --- | --- | --- | --- | --- | --- |
| A | .10<br>(.113) | .007<br>(.007) | .010<br>(.007) | .032<br>(.032) | .055<br>(.050) | .080<br>(.068) | .017<br>(.018) | .031<br>(.041) | .036<br>(.029) |
| B | 0.997 | .101<br>(.106) | .010<br>(.007) | .036<br>(.032) | .055<br>(.050) | .079<br>(.066) | .019<br>(.018) | .031<br>(.041) | .037<br>(.029) |
| C | 0.996 | 0.997 | .101<br>(.104) | .040<br>(.035) | .056<br>(.052) | .076<br>(.064) | .017<br>(.017) | .030<br>(.039) | .039<br>(.030) |
| D | 0.966 | 0.955 | 0.947 | .109<br>(.130) | .044<br>(.039) | .095<br>(.074) | .041<br>(.039) | .052<br>(.061) | .034<br>(.030) |
| E | 0.946 | 0.941 | 0.941 | 0.952 | .139<br>(.155) | .072<br>(.063) | .056<br>(.050) | .061<br>(.071) | .058<br>(.051) |
| F | 0.942 | 0.948 | 0.961 | 0.873 | 0.922 | .169<br>(.175) | .065<br>(.059) | .080<br>(.076) | .094<br>(.077) |
| G | 0.995 | 0.993 | 0.993 | 0.946 | 0.932 | 0.956 | .112<br>(.121) | .032<br>(.044) | .045<br>(.042) |
| H | 0.929 | 0.932 | 0.937 | 0.859 | 0.878 | 0.906 | 0.932 | .103<br>(.104) | .053<br>(.051) |
| I | 0.961 | 0.957 | 0.956 | 0.960 | 0.941 | 0.936 | 0.953 | 0.877 | .092<br>(.101) |

\*A: main specification as reported in the main text
B: main specification + additional weather variables (Table S10 column 1)
C: main specification + absolute humidity (Table S10 column 2)
D: main specification with top 5% $R_0$ excluded
E: main specification with first 15 days excluded (Table S5 column 2)
F: main specification with US-only sample
G: main specification with top 5% UV index excluded (Table S8 column 3)
H: main specification + interaction between UV index and precipitation (Table S10 column 4)
I: main specification with location-specific quadratic trend (Table S4 column 5)

### 5. Specification and validation using synthetic data

We conducted extensive synthetic analyses to inform the selection of a reliable statistical method and to build confidence in our final estimation method. These analyses could be divided into those focused on specifying the estimation method (5.2), and those designed to validate our method in a study wherein the analyst was blinded to the true specification (5.3). After providing an overview in 5.1, we explain these two sets of analyses. Detailed codes are available at <https://github.com/marichig/weather-conditions-COVID19/>.

#### 5.1. Summary of our approach and findings

Before providing the details of the analysis, we review the main objectives, approach and findings of this test. We then provide more details about the analysis in sections 5.2 and 5.3.

- **Approach:** To build and validate our method and examine its sensitivity to different assumptions, we created synthetic data from simulated epidemics with several different assumed temperature effects on infection. The true exposure, exact detection delays, and temperature functions were hidden from our estimation method to objectively assess the method’s success. Our objectives were two-fold and we created a task allocation among researchers to meet those objectives. First, investigator HR used the iterations of this process to improve our statistical estimation method and ensure that it was able to find temperature functions under various assumptions (Section 5.2.2). Second, we used a more realistic individual-level model of infection (stochastic agent-based model) built by two other investigators (NG and MG) not involved in the first synthetic data analysis (used for method design) to assess if our statistician (RX), who was unaware of true functional forms or the second model structure, could correctly identify effects in this different simulation environment (section 5.3). This design addressed the risk that a method fine-tuned on synthetic data may perform well under the assumed simulation setting but fail in other environments.
- **Results:** The key finding from these experiments (further elaborated below) include: 1) Our preferred model specification could accurately identify correct weather impacts if true infection was observable; 2) In the absence of data on true exposure (which is the case in COVID-19 due to testing delays), however, estimation of weather impact becomes complicated, and many intuitive specifications used in other studies fail to recover true impacts. This challenge may afflict many attempts to identify the link between weather and COVID-19 transmission; 3) Our algorithm for uncovering the true exposure and the specification we selected offer a potentially conservative but qualitatively informative view of the true underlying impacts; 4) Our preferred specification is robust to a few key uncertainties that may vary between simulated numbers and the actual epidemic; and 5) A statistician blinded to true data generating process was able to use this method to identify true weather effects from synthetic data generated from a different, more detailed, agent-based model of COVID-19 epidemics. Given these results from the analysis of the synthetic data, we can have more confidence in the analysis using the actual data.

#### 5.2. Statistical specification using synthetic data

Our approach consists of building a simulation model of the epidemic to generate synthetic data (with known weather impact functions) followed by estimating various statistical specifications to assess their ability in identifying the true functional forms.

##### 5.2.1. Simulation model

We used a simple SIR-based simulation model to generate synthetic epidemics. This model was applied across various locations (with different vectors of weather variables (*W*(*t*)) impacting epidemic curve based on *g*(*W*(*t*))) to generate the raw data going into alternative statistical methods to identify the function *g* below. Equations of the simulation model are presented in Table S12.

**Table S12.** Equations of the SIR-based stochastic simulation model of epidemics Iterates daily until epidemic ends or until t=50 is reached.

|  |  |
| --- | --- |
| $I_N(t) = \text{Poisson}\left(\frac{bS(t)C_I(t)}{S(0)}p(t)g(W(t))\right)$ | New infections assumed Poisson based on susceptible stock ($S$), Infectious stock ($C_I$), force of infection ($b$), impact of distancing policies ($p(t)$), and weather effect ($g(W(t))$). We focused on mean temperature as the variable going into the $g$ function. |
| $p(t) = \text{Min}(1, e^{-s(t-t_0)})$ | Distancing policies reduce risk of infection by a fixed fraction ($s$) per day, beginning the first day with a confirmed case ($t_0$). Parameter $s$ is drawn uniformly from 0.03-0.05 range. |
| $S(t+1) = S(t) - I_N(t)$ | Updating susceptible stock for next period |
| $R(t + 1:t + 21) +$<br>$= Multinomial(I_N(t), D_{Rcv})$ | Adding to future daily recovery rates ($R$) to incorporate the recovery of all those infected today using a multinomial distribution. $D_{Rcv}$ is the recovery delay distribution, assumed Poisson with mean of 20 days. The “+=” operator adds to the existing vector on the left-hand side values on the right hand. |
| $I_M(t + 1:t + 18) +$<br>$= Multinomial(round(I_N(t)$<br>$* f), D_{Dtc})$ | Adding to future measurement of detected cases ($I_M$) to incorporate fraction $f$ of those infected today in future detection data. A multinomial distribution is used following the Detection Delay lag structure ($D_{Dtc}$; see section 2 of Appendix). In our baseline model we use $f = 0.1$, and test more complex functional forms where $f$ increases over time from zero to a maximum of 0.3 in response to measured infections in 3 different scenarios. |
| $C_I(t + 1) = C_I(t) + I_N(t) - R(t)$ | Updating next period stock of infectious based on recovery and new infections. Initial infectious population of 3 is assumed so that few epidemics die out due to stochasticity. |
| **Initialization and functional assumptions** |  |
| $S(0)$<br>$= Max(1000, Normal(5e5, 2e5))$ | For different simulated locations we initialize the susceptible population to vary a wide range. In lieu of explicit response mechanisms, these different sizes represent different slow down trajectories in the new infections over time. |
| $b = \frac{4}{20} Normal(1, 0.3) h(W)$ | For different simulated locations the baseline force of infection is drawn from a normal distribution, set to have an expected basic reproduction number of 4, and to be potentially correlated with weather variables (using $h(W)$ term) to ensure robustness of results to such correlations. Note that parameter $b$ is a daily factor, and thus divided by 20 to capture the expected duration of illness. |
| $g(W)$ | We use different $g$ functions, including quadratic and linear forms. We focus on temperature as the primary $W$ variable that is read from data and input into the synthetic data. |
| $h(W)$ | In our main synthetic analysis, we use:<br>$h(W) = \frac{40 + Mean(W(1:50))}{50}$<br>This creates a positive correlation of 0.47 between temperature that affects transmission rates and the reproduction number for a specific location. |
| $W$ | We use mean daily temperature data from a start date uniformly picked between January 23<sup>rd</sup> 2019 and March 25<sup>th</sup> 2019 for $t=1$ for each location, and continue accordingly. 2019, rather than 2020, temperature data are used to ensure sufficient data are available for full epidemic to unfold. |

Using these specifications, we then conducted multiple experiments to identify a viable specification. In each experiment we simulated the model for 20 iterations (with different random realizations) for a sample of 500 locations randomly drawn from our 3,739 locations with their actual temperature data. Temperature data started from a random day between January and March, and then were fed into *W*.

We summarize the results from each experiment using a graph of the shape of the estimated relationship between temperature and natural logarithm of estimated reproduction number, as we do in our main statistical analysis. This estimate is then compared with the true relationship (*ln*(*g*(*w*)); the thick dashed line in figures below). The success measure for our method is to have the estimated relationship from our method close to the thick dashed line. Effects falling between the true curve and the horizontal line at zero would be conservative, and those falling outside this range may be misleading. Our actual temperature data in the simulation period are bounded to smaller ranges (90% of data falls between −10 and 20 degree Celsius) than reported in these figures. Therefore, extrapolations outside this range are not necessarily indicated by the data, rather, emerge from the estimated functional forms. Nevertheless, we graph a much wider temperature range (−30 to 50) to highlight the errors of such extrapolation. We also report the means of estimated parameters for *ln*(*g*(*w*)), 95% confidence interval, coefficient of determination (r^2^), and sample size for each experiment.

##### 5.2.2. Synthetic experiments

In the rest of this section, unless specified in the top row of a figure, we focus on quadratic equations for *ln*(*g*(*w*)). The main specification uses *ln*(*g*(*w*)) = −0.01w − 0.002*w*^2^. We also conduct sensitivity analysis to other functional forms for *ln*(*g*(*w*)). We summarize the results of these synthetic analyses under 10 experiments, which could be categorized in three subsets. Experiments 1 to 3 (sections E1–E3 below) introduce the main challenges in correctly associating weather with reproduction number, concluding that while not an impossible task, the best one can expect from similar efforts may be to find estimates that are conservative but not misleading. Next, in experiments E4 to E6 we show support for the chosen statistical specification against other plausible alternatives. Finally, experiments E7 to E10 show the robustness of the preferred specification to a variety of assumptions.

###### E1) Endowed with true, deterministic infections, the method finds the correct impact of weather. Relaxing either assumption deteriorates results

In Figure S5, we compare three different scenarios. In the first two (A and B), the estimation method is provided with the actual true infections (rather than those estimated using the method discussed above). Moreover, the first experiment (A) also assumed deterministic infection rate (that is, *I_N_*(*t*) *= E*(*I_N_*(*t*)) in Table S12). Plot C shows results using our baseline specification: estimating true infections using quadratic programming, including location-specific trend lines and fixed effects, dropping days with estimated exposure below 1, as well as the first 20 days after the estimated exposure first reaches 1, and excluding the outlier estimated reproduction numbers (those above 95%). The two assumptions on using deterministic infection and true infections in the first two experiments are not realistic. Instead, they reassure us that the method, given correct exposures, would find the true functional forms. Moreover, they inform the challenges to unbiased estimation of reproduction number due to stochasticity of infections (comparing plots A and B) and estimation of true infections from reported data (comparing plots B and C).

Inspection of these results reveals two major challenges to estimating reproduction number: i) Randomness in infection rate leads to weaker identified effects. ii) The imperfect identification of true infections from reported cases significantly reduces the magnitude of estimated effects. Both of these effects generate a bias towards null estimated effects, even when true effects are very significant. As the experiments reported in the following sections show, our baseline model, despite its conservative estimates, might be among the best available options to find estimates for the impact of weather on transmission rates.

Note that the true linear and square terms in *ln*(*g*(*w*)) are reported in the title for each panel in the following figures.

**Figure S5:**
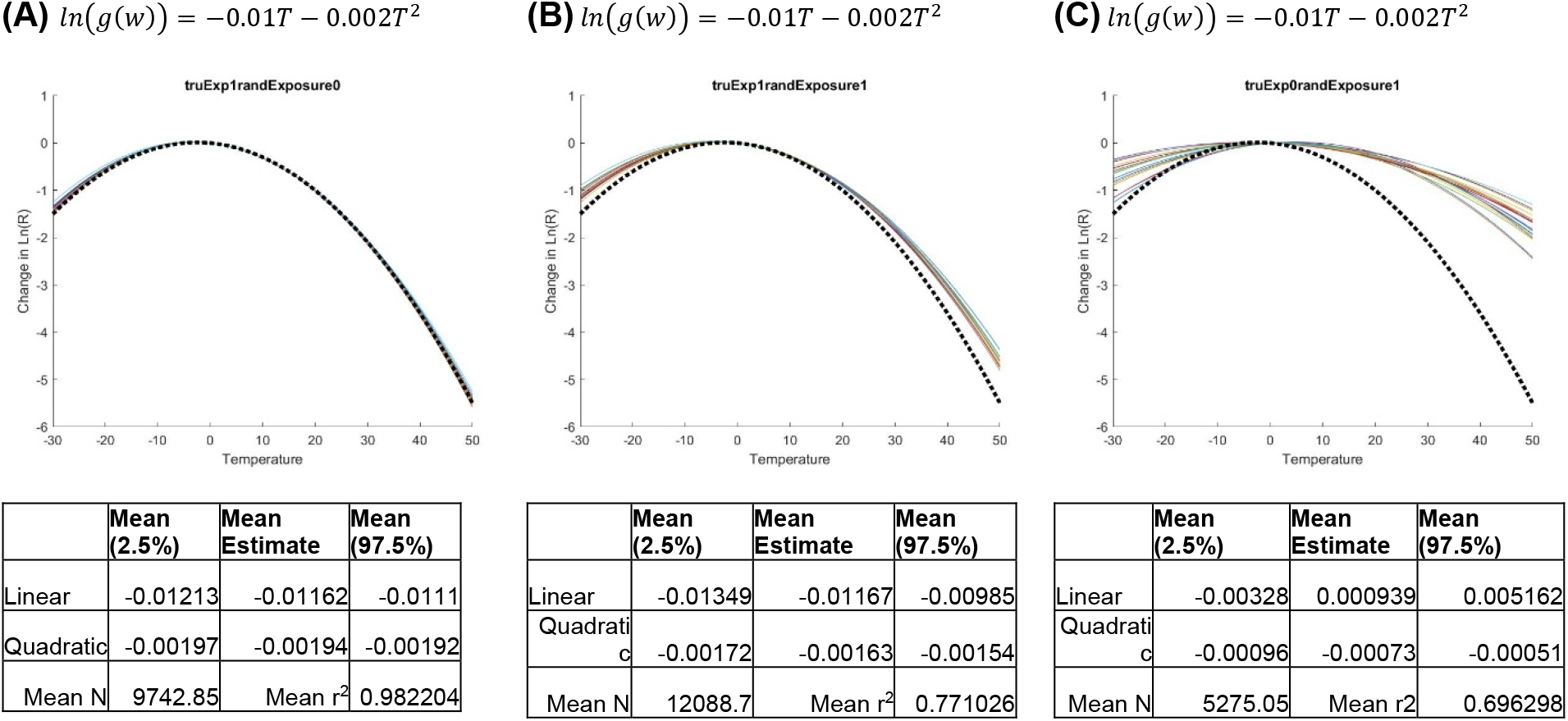
Impact of stochasticity in infections and imperfect estimation of true infections on quality of estimated parameters.

###### E2) Using true infections offers reliable, and slightly conservative, estimates for a range of functions

Before focusing on the main specification, we report another set of experiments that show the performance of estimation method with true infections under three other functional forms (Figure S6). The main observation in this set of experiments is that true infection rates, even including randomness in infection, would offer close estimates for the underlying impacts of temperature on reproduction number across a range of functional forms. So, the statistical method in use is fundamentally sound.

**Figure S6:**
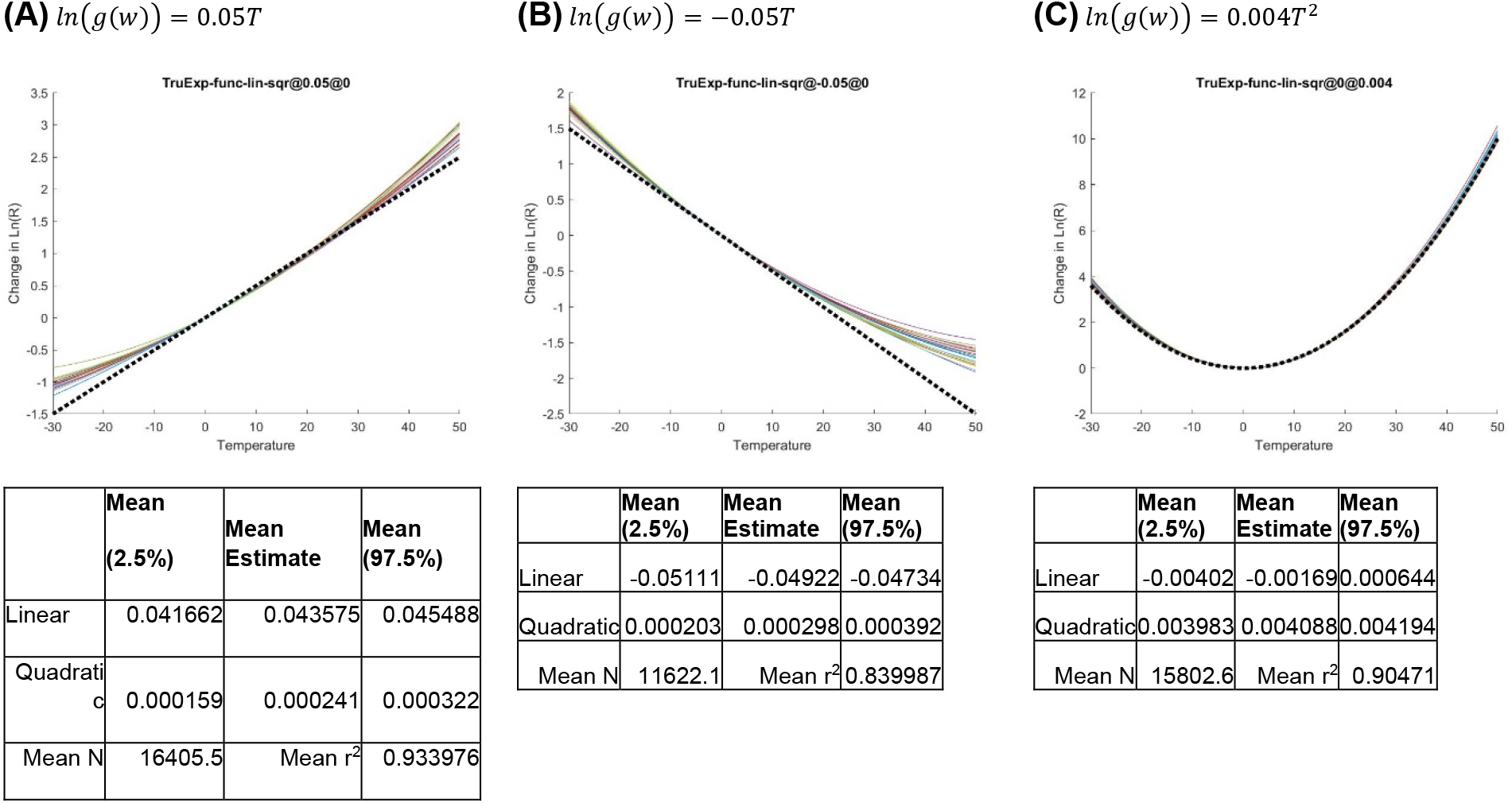
Performance of estimation method in identifying different functional relationships between weather and reproduction number when provided with true infections.

###### E3) Using proposed estimation method provides conservative, but largely consistent, estimates

In the next set of experiments, we test the main estimation method, which uses estimated exposures, to find the effect of temperature under the different functional forms introduced in E2. In these experiments, we continue to use the same inclusion criteria used in experiment 1. Overall, estimated effects, as shown in Figure S7, are qualitatively consistent with the true functional forms, but also show important deviations: i) The results include some biases in estimated parameters when the estimated functional form differs from the true function (e.g., including quadratic terms that are not in the true function). This is a general feature of estimating mis-specified functions. ii) Results are generally conservative (pointing towards null effects) in the regions of the temperature actually covered by W data. Based on these observations, the use of the estimation method should include appropriate caution. The results may undervalue the true magnitudes of weather effects, and if more complex functional forms are estimated, spurious results may be found. For this reason, in our main specification we limit the use of more complex interaction terms.

**Figure S7:**
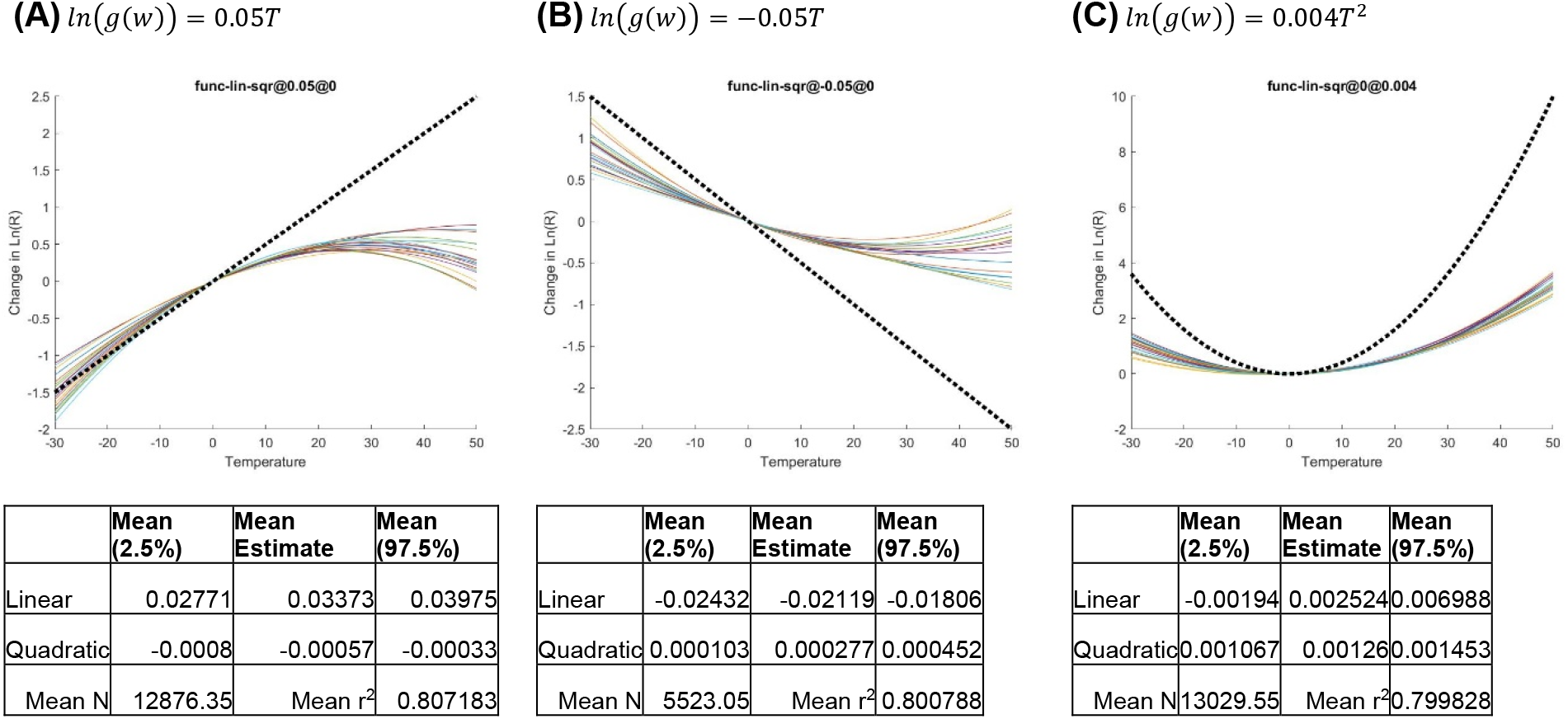
Performance of preferred estimation method with realistically available data in identifying different functional relationships between weather and reproduction number.

###### E4) Results with simple shifting of infections

Our preferred specification uses the estimated infections based on the quadratic programming method discussed in section S3. Here we compare those results against a simpler specification that shifts back detected infections each day by 10 days to infer the true infection rate on each day. Results are shown in Figure S8. We assess this alternative under the same functional forms discussed in experiment 3 (E3) and thus results are directly comparable with that experiment. In short, the simple shift method offers results that are comparable with the preferred specification but more conservative (e.g., Panel C) and in some cases more biased (e.g., stronger, incorrect, quadratic term in Panel A). In a few other experiments with other simulation model setups, we found that this intuitive specification (simple shifting) may significantly underperform our calculated exposure method when behavioral responses are more complex.

**Figure S8:**
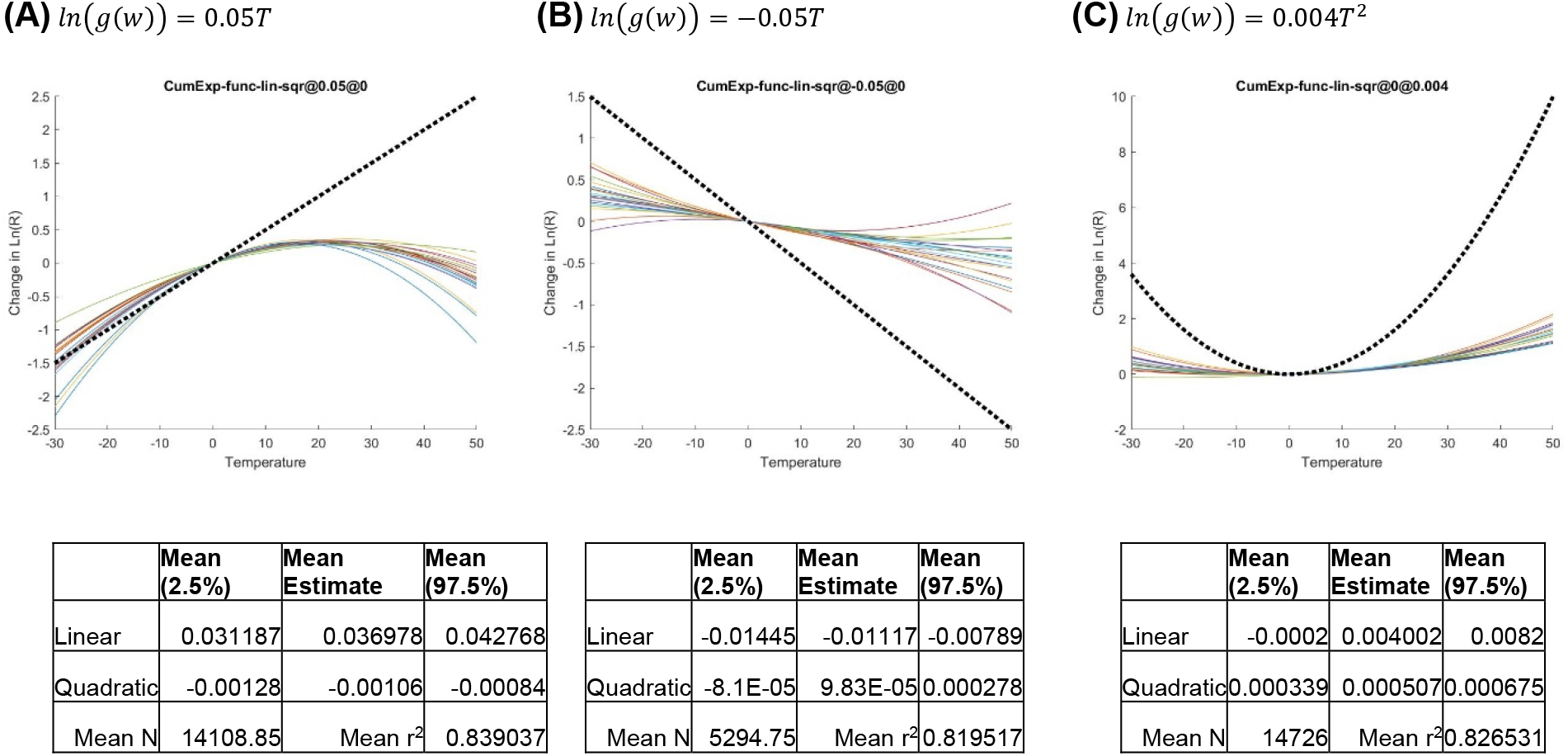
Impact of using a simple shift of measured infections on performance of estimation under various functional forms.

###### E5) Using other combinations of location specific effects

In this experiment, we compare three settings in which we exclude fixed effects but keep location specific trends (Figure S9, panel A), exclude trends but include fixed effects (panel B), and exclude both fixed effects and trends (panel C). These experiments are directly comparable with our preferred specification where both trends and fixed effects are included (Panel C in Figure S5).

The overall performance deteriorates significantly when location specific trends are removed (e.g., panels B or C compared with baseline results). This should be expected; the positive trend in temperature during the spread of epidemic in late winter and early spring likely correlates with behavioral and other responses that temper the spread in each location. Thus, excluding location trends would lead regression results to pick up that spurious correlation and inflate the impact of temperature, creating illusory and misleading results. Thus, including location specific trends is a necessity.

The results of excluding fixed effects but including location-specific trends are comparable with our baseline findings and somewhat stronger (closer to true effects) in this experiment (but also in some other experiments not reported here). Therefore, one could argue for inclusion of only location-specific trends rather than both fixed effects and location-specific ones. The theoretical logic for such recommendation is that two different location-specific parameters absorb much of the variations in weather between and within locations, and combined with errors in the identification of true infections from reported data, very modest signal remains to estimate the weather function leading to weak coefficients.

The risk with excluding fixed effects is that we would not be able to provide appropriate controls for a host of unobserved location-specific characteristics, from cultural norms in interaction and eating, to public transportation use, comorbidities, and age distribution, which may conceivably interact with transmission *and* be correlated with some of our weather and pollution variables (thus introducing unknown biases). Moreover, R-squared drops substantially in panel A in comparison to the baseline of Panel C in Figure S5 (with fixed effect and location-specific trend effect), showing that there are cross-regional variations missed if we don’t control for fixed-effect variation. We therefore decided to select the more conservative specification (with both fixed effects and location specific trends) as our primary model.

**Figure S9:**
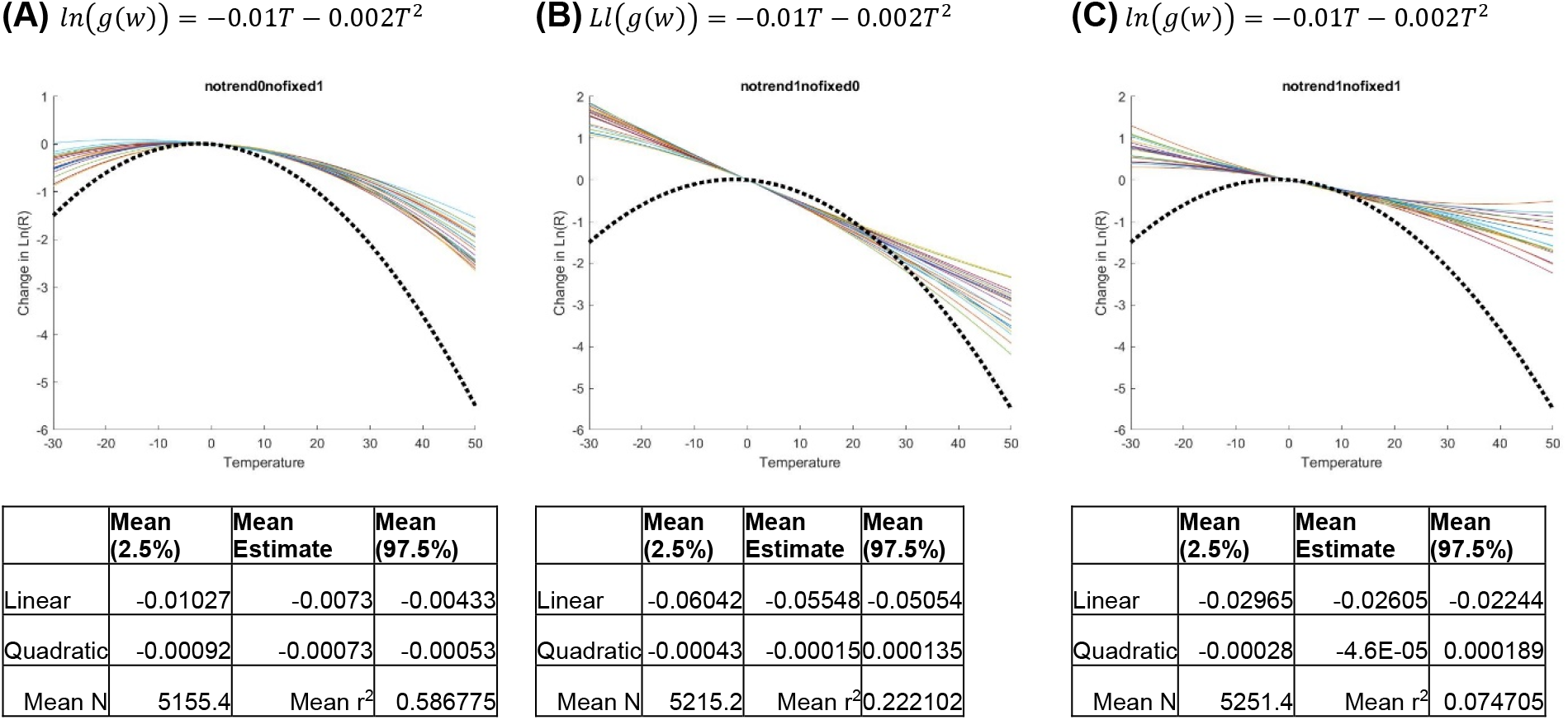
Comparing the use of only trend effects (A), only fixed effects without trends (B), and no location specific fixed or trend effects (C).

###### E6) Weighting data points in regression do not improve performance

One potential issue in the current specification is that locations with larger outbreaks are weighted the same way as locations with smaller outbreaks. The data from larger outbreaks may well be more reliable, and the estimates of R calculated from that data thus more reliable. We assess if a correction for this issue can improve estimation results. To do so we use simulations to estimate how the variance in the dependent variable *ln*(*R*) scales with the number of estimated daily true infections and use that estimated variance to conduct weighted least square regressions. Results, reported in Figure S10-panel A (panel B showing baseline replicated from experiment 1), show more conservative outcomes and dispersion compared to the unweighted regressions and no significant improvements. Hence, we do not pursue this correction in our main specification.

**Figure S10:**
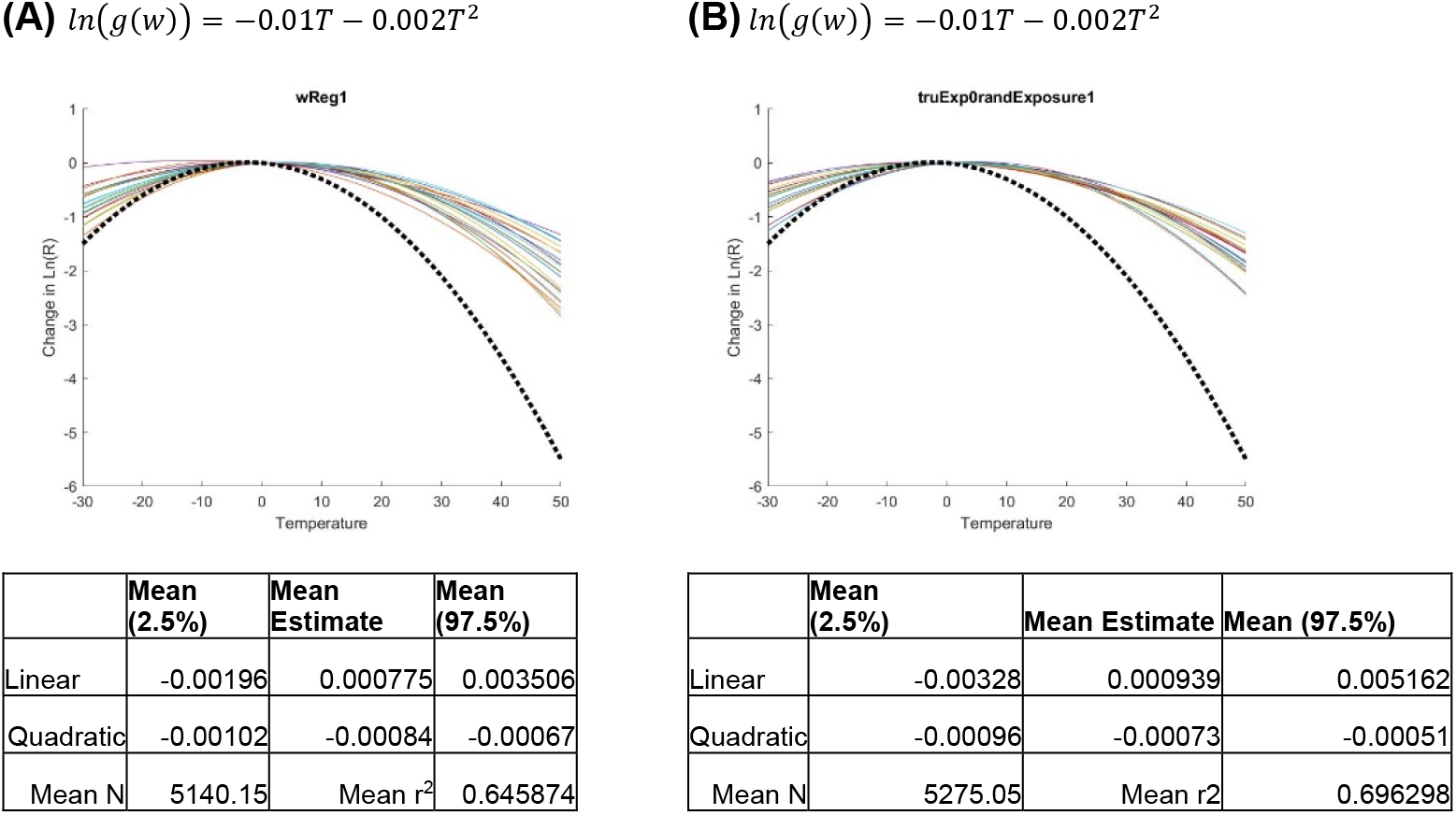
Impact of using weighted regression (A) vs. unweighted (B)

###### E7) Results are robust to variance in susceptible population

In the baseline simulations, the location-specific population size, which partially controls the speed of spread, is normally distributed with mean of 500,000 and standard deviation of 200,000 (with 1,000 population minimum). In Figure S11, we compare that baseline (panel B) against standard deviation of 0 and 400,000. The impacts are largely negligible, suggesting that the variance in speed by which the spread slows down due to herd immunity does not impact the findings much. This is largely expected, as in most simulated epidemics the transmission halts as a result of behavioral response and not herd immunity dynamics.

**Figure S11:**
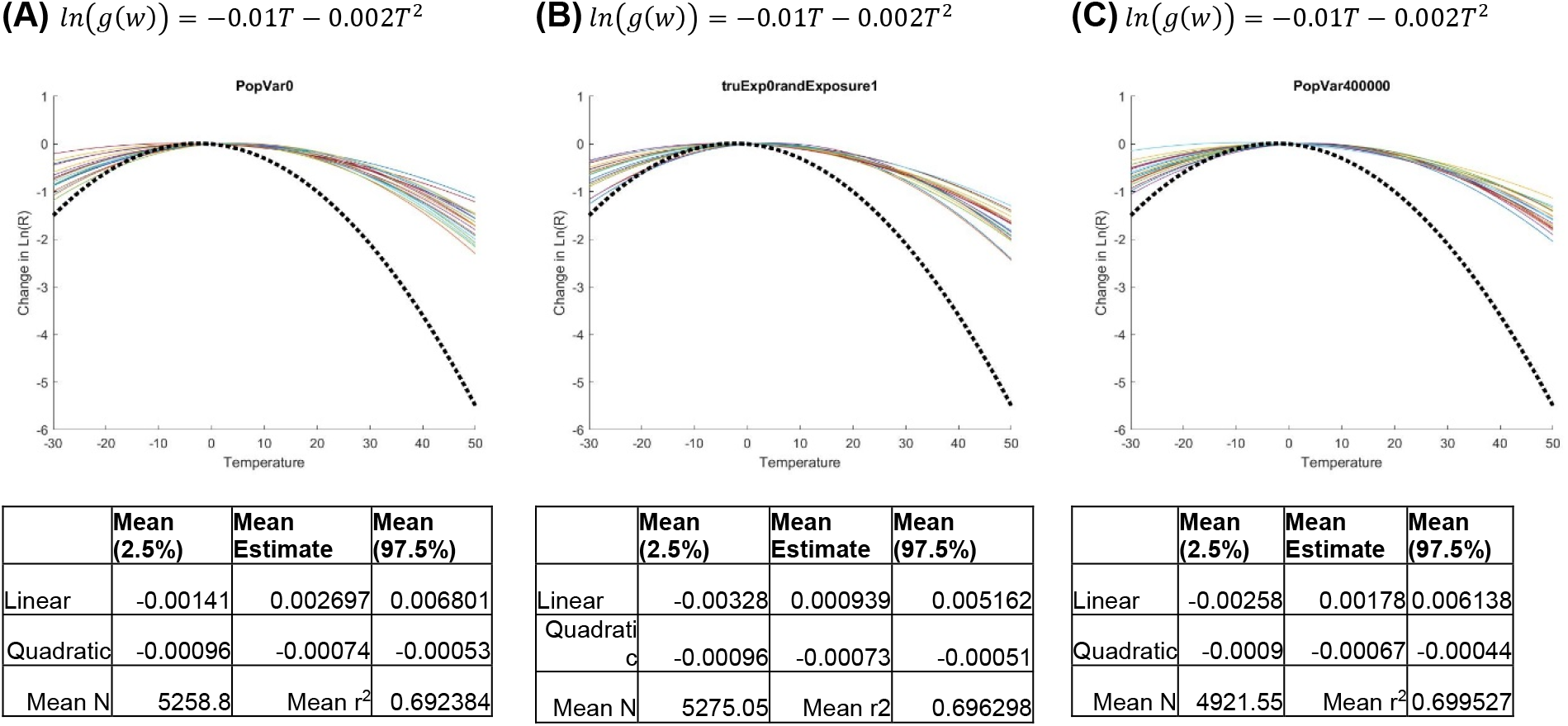
Impact of various variances in population size per location. (A) No variance (mean is 1e5). (B) Baseline (Standard deviation of 200,000). (C) Standard deviation of 400,000.

###### E8) Variations in basic reproduction number do not impact the findings

In the baseline simulations (E1), the location-specific basic reproduction number had a mean of 4 and variance of 1.2 (normally distributed with a minimum of 0; but also positively correlated with average temperature in the location). In Figure S12, we compare that baseline (reproduced in panel B) against standard deviations of 0 (Panel A) and 2 (Panel C) across locations. The impacts are largely negligible, suggesting that the variance in speed by which the spread grows does not impact the findings.

**Figure S12:**
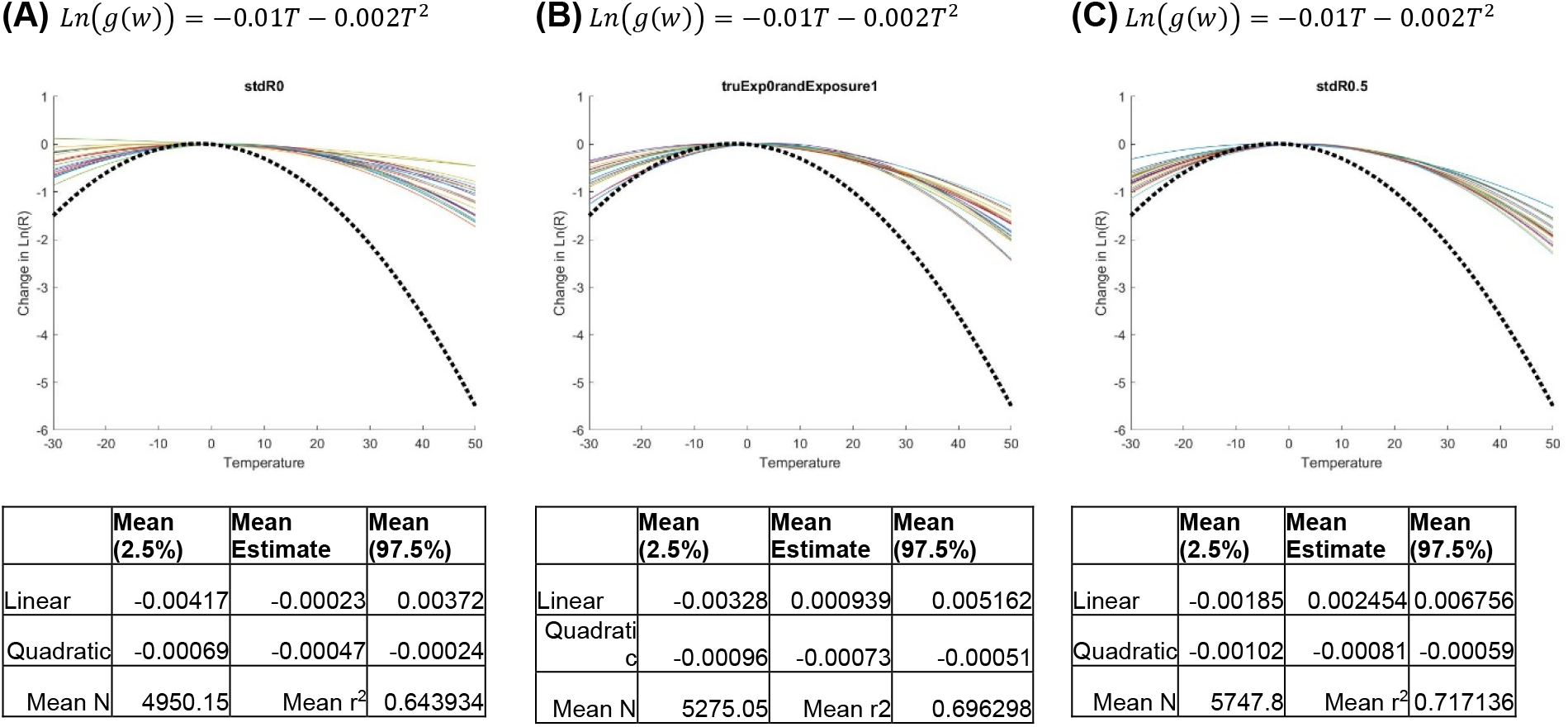
Impact of different variances in basic reproduction number. (A) Standard deviation of basic reproduction number is 0, with mean of 4. (B) Standard deviation of basic reproduction number is 1.2. (C) Standard deviation of basic reproduction number is 2.

###### E9) Correlation between basic reproduction rate and temperature has limited impact on results

Another variant on the distribution of basic reproduction number considers its correlation with the temperatures informing the weather function. In the baseline specification and all the experiments so far, we used a correlated version of that relationship (with a correlation of 0.47 between basic reproduction number and average temperature; see simulation model specification in Section 5.2.1). Here we compare that setup (reproduced in Figure S13, Panel B) with the uncorrelated version where basic reproduction number is independently drawn for each location with a mean of 4 and standard deviation of 1.2 (Panel A). Results have limited sensitivity to this potential correlation.

**Figure S13:**
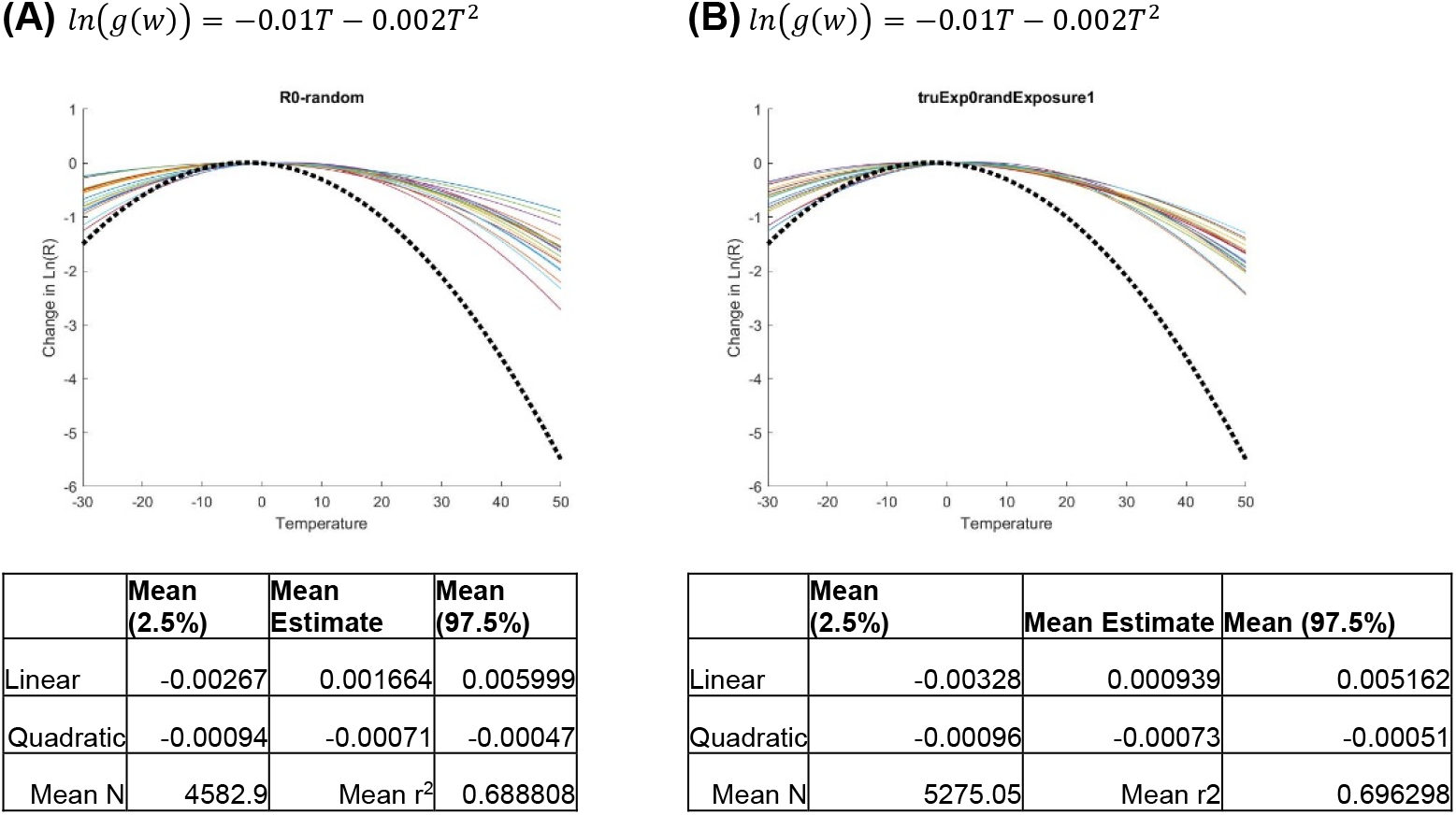
Impact of including correlation between basic reproduction number and average location-specific temperature (baseline; reproduced in Panel B) vs. having no correlation (panel A).

###### E10) Changes in test coverage do not change the results

The simulation model so far assumed a constant test fraction of *f* = 0.1, that is, in expectation only 10% of infections were detected. This ratio may change over time as test capacity ramps up in response to infection measures in practice. Here, we explore results under three such ramp-up scenarios. In Figure S14 panels A-C, the following ramp-up scenarios are assumed as a function of confirmed infections (*I_M_*): A) *f = Min*(0.2,0.001*I_M_*); B) *f* = *Min*(0.2,0.05Log_10_(*I_M_* + 1)); and C) *f = Min*(0.2,0.01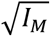). Overall, results are rather insensitive to these very different test fraction numbers, suggesting robustness to this consideration.

**Figure S14:**
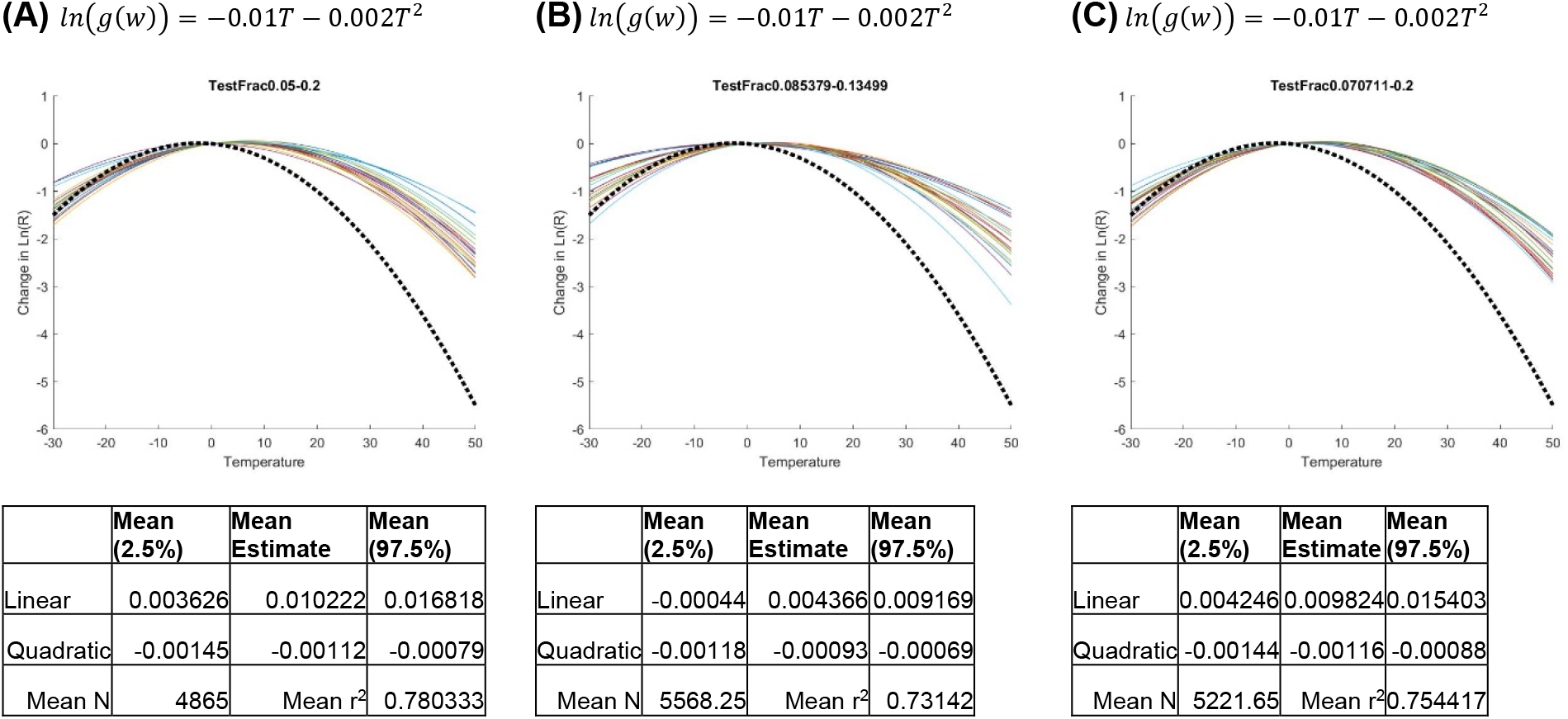
Impact of three different test coverage functions.

###### 5.2.2.4. Summary of Synthetic Experiments

The key finding from these experiments include: 1) The model specification we use could identify correct weather impacts if true infection were observable; 2) In the absence of that data estimation of weather impact becomes complicated, and many intuitive specifications fail to recover true impacts. This may be a general challenge afflicting any attempt to identify the link between weather and COVID-19 transmission; 3) The specification we selected offers a conservative but qualitatively sound view of the true underlying impacts. For example, in most estimations the quadratic term is estimated at about half its true value; and 4) This specification is largely robust to the key uncertainties that may vary between simulated numbers and the actual epidemic.

### 5.3. Blinded study specification and results

In this last step of synthetic data analysis, our objective was to use a more detailed individual-level model of infection (stochastic agent-based model) of interacting individuals to create synthetic data of reported cases, distort the outcome with a delay function to represent test/report, and examine whether our statistical methods are still capable of finding our weather functions. Two of our co-authors (NG and MG) created synthetic data and hid their assumed temperature effect functions from our statistician (RX), whose task was to discover the assumed temperature effect function.

To that end, we created an agent-based simulation model of infection and simulated the model for 100 hypothetical towns of different populations, different *R_0_*s (potentially due to different contact rates and population density), different start days of infection, and different temperatures. We started from the generic individual model of infection (available on the NetLogo library) that is consistent with the basic SIR model at individual level. We modified the model using parameter values that are more consistent with COVID-19, and included several features needed to import and export data to the model. We modified the infection function to include the temperature effect on the probability of infection. We used three major scenarios for temperature effect (inverse U-shaped effect, linear increasing effect, and no effect (placebo)). The scenarios included actual temperature values coming from a sample of 100 regions from the real-world data. The ABM model’s output was generated using a detection delay with Poisson distribution with mean of 10 days. These data were used to estimate true infections with the method discussed in section 3. The model codes are available at https://github.com/marichig/weather-conditions-COVID19/. Figure S15 shows an example of creating synthetic data (scenario 1, explained in the following) with Panel A showing the true cumulative infections and panel B showing the reported values.

**Figure S15:**
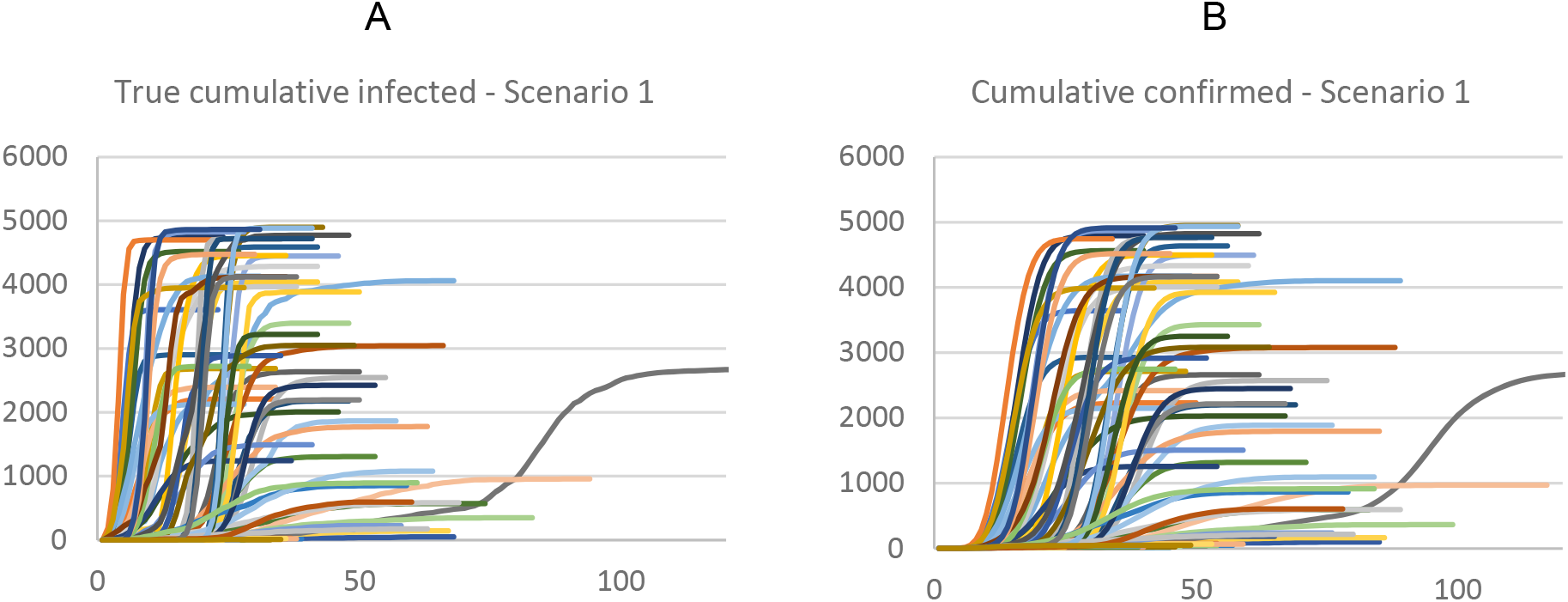
An example of the synthetic data generation process with an assumed temperature effect function hidden from our statistician. True cumulative cases (A) and cumulative number of confirmed cases (B).

The tests (scenarios) included quadratic (S1), no effect (S2), and positive linear effect (S3). For all scenarios we tested models both including fixed and trend effects (Si1) and those with only trend effects (Si2). In non-fixed-effect tests, in order to make a control variable consistent with our main regression, we added one extra variable, a hypothetical variable of “population density,” to represent variations in locations correlated with basic reproduction number. In this setup, population density was correlated with the basic reproduction number excluding temperature-related factors (*ρ* = .8). Our statistician did not know the true temperature function, so he used both linear and quadratic terms to map the predicted temperature effect in all cases, even when the effect was linear.

Results are graphically summarized in Figure S16; for each scenario the results are compared with the “true” function of temperature (darker lines). Overall our statistician was able to correctly estimate the sign and magnitude of temperature effect in all cases, while the effect was generally underestimated further supporting the proposition that the method offers conservative estimates (e.g., in Figure S16, left panel, compare S11, and S12 curves with the true effect of “S1-true effect”). Also, in line with the previous section, we find that including fixed effects may lead to somewhat more conservative estimates than excluding it, but results are less prone to other biases. Higher R-squared values were obtained with fixed-effect models.

**Figure S16:**
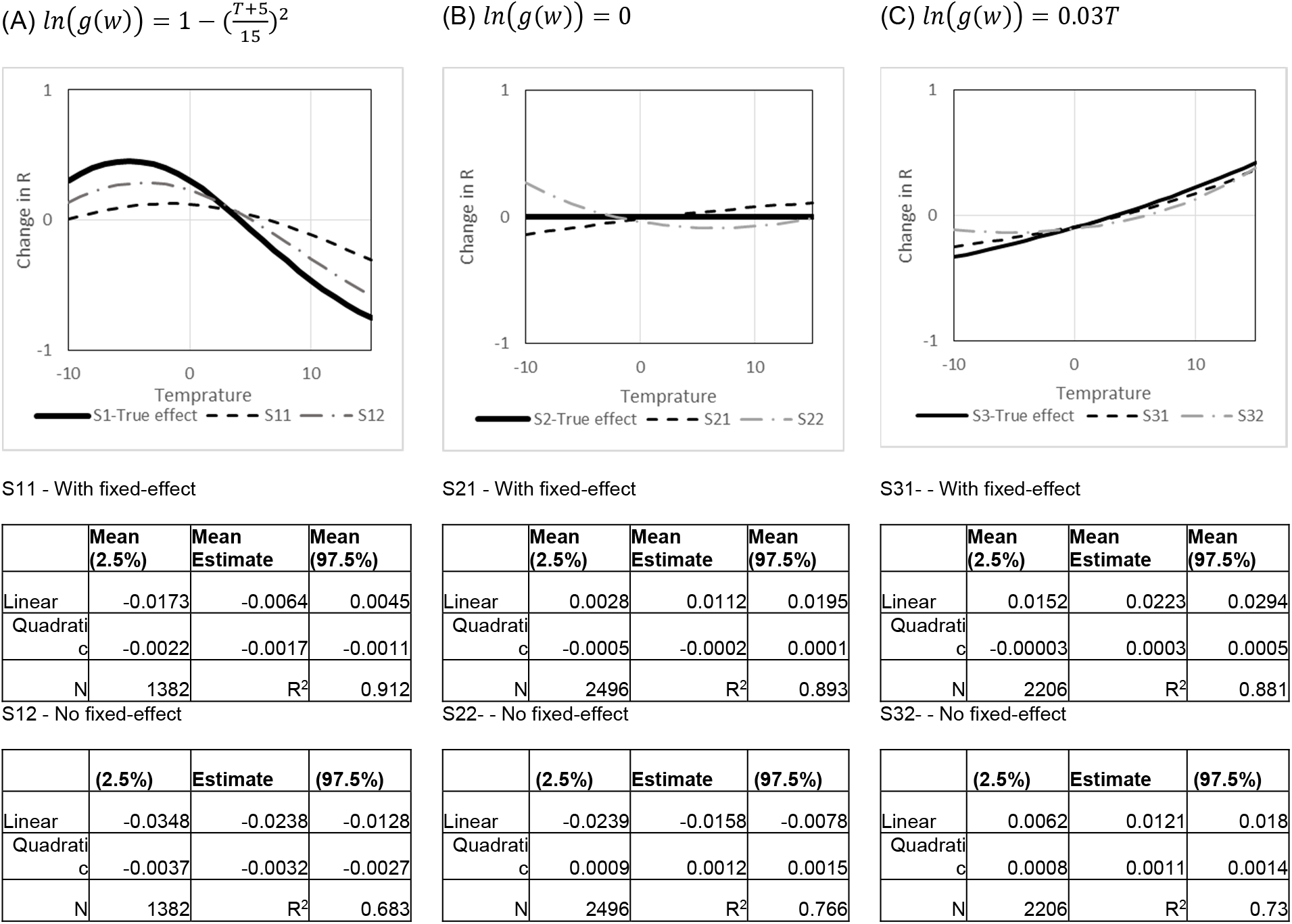
A comparison of assumed functions in the agent-based model (dark lines) and the outcome of our regression analysis (dashed lines). Y-axis is change in R in comparison to average R. Note: S_ij_ represents results from Scenario *i* under conditions of fixed-effect (j=1), and no fixed effect regressions (j=2).

## 6. Global projections over time

In this section we report graphs of projected Covid-19 Risk Due to Weather (CRW) at 4 different time periods in the coming year (Figure S17–Figure S20). These projections use a 15-day moving window to average different weather and pollution variables in the previous year (2019-2020) (for weather) or 2019 (for pollution, since pollution in 2020 is affected by COVID-19 related behavioral changes) and use those averages as the predictor for the coming year (2020-2021). Daily projections year-round for these global locations, the largest 1072 cities across the world, and US counties are available at: https://projects.iq.harvard.edu/covid19.

The main drivers of changes observed include temperature and UV effects, which in some regions and times of the year pull the estimates in opposite directions (with U-shaped effect for UV, high values of UV lead to more transmission while that is also often correlated with higher temperature). This interaction creates some of the higher risks in summer for the regions close to the equator.

**Figure S17:**
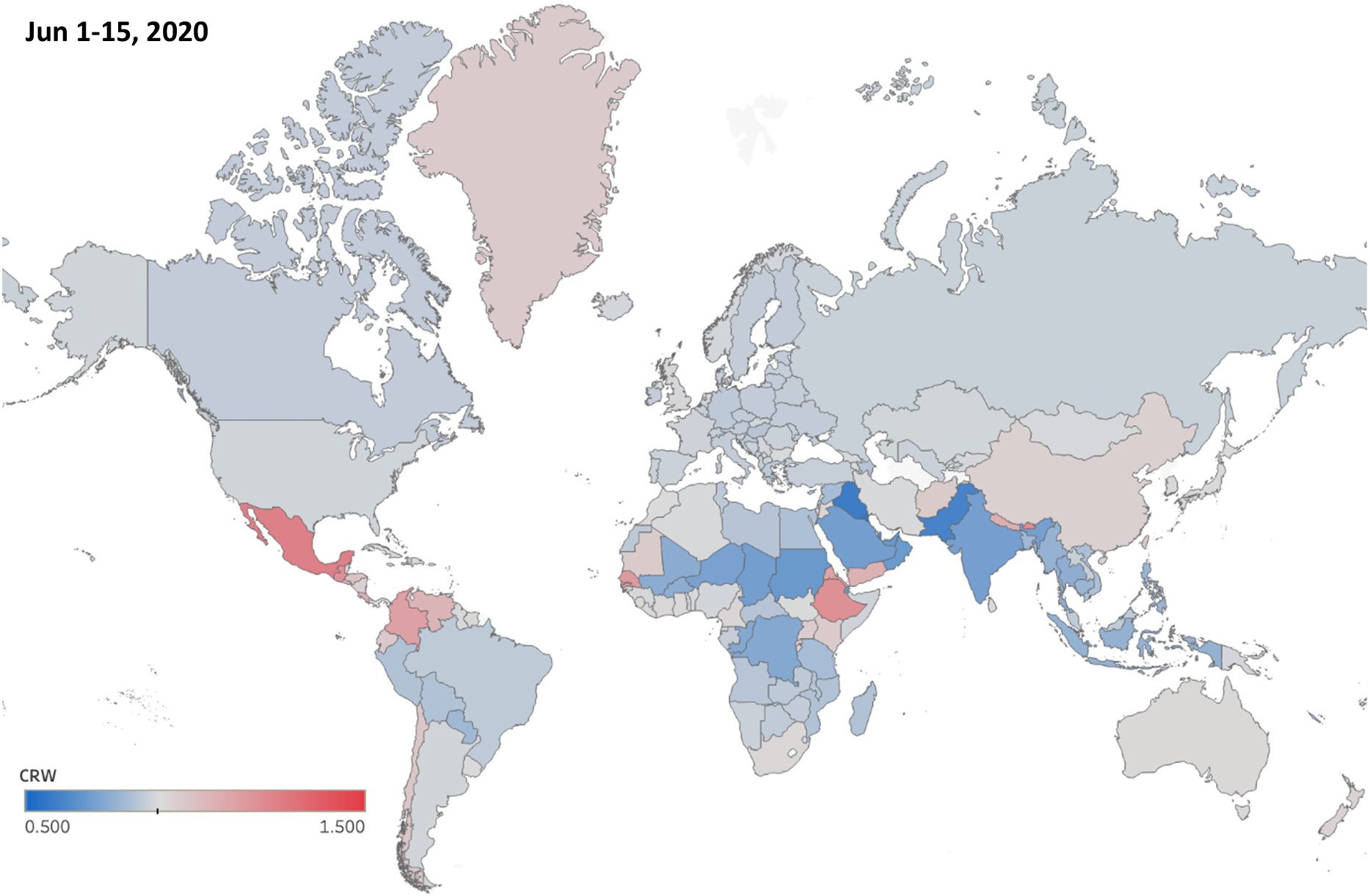
Projected Relative COVID-19 Risk Due to Weather (CRW), averaged for the first half of June 2020

**Figure S18:**
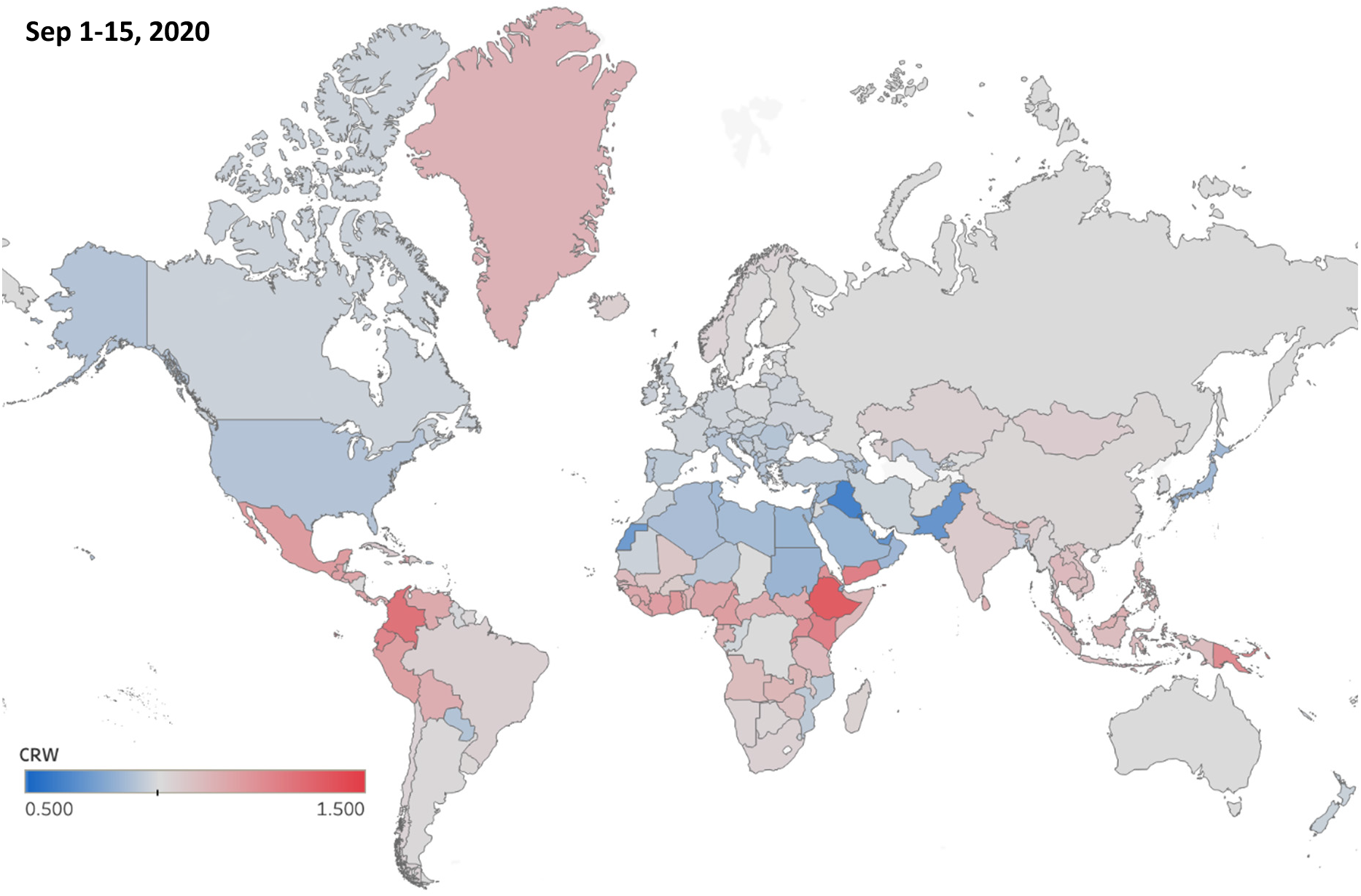
Projected Relative COVID-19 Risk Due to Weather (CRW), averaged for the first half of September 2020

**Figure S19:**
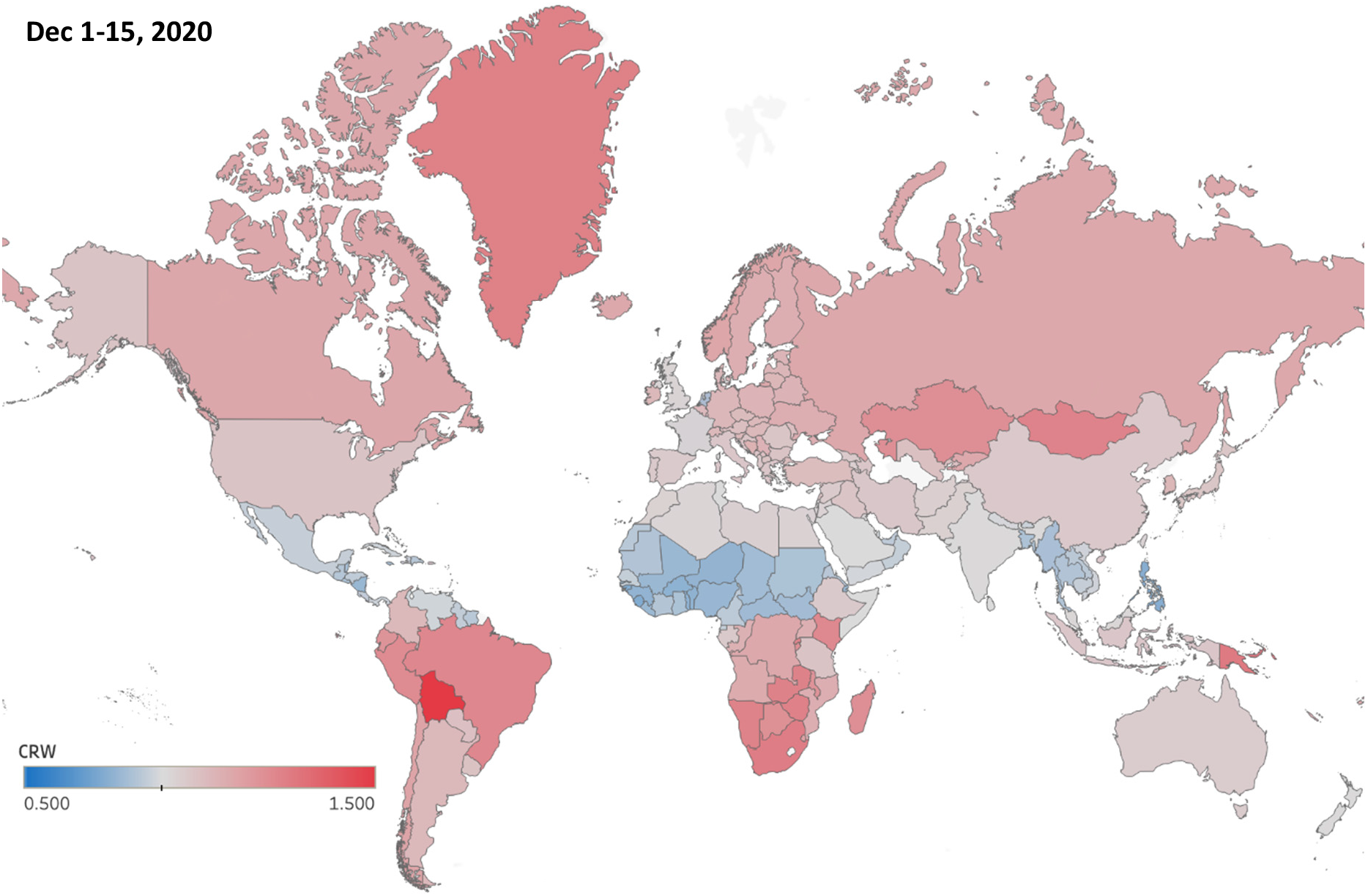
Projected Relative COVID-19 Risk Due to Weather (CRW), averaged for the first half of December 2020

**Figure S20:**
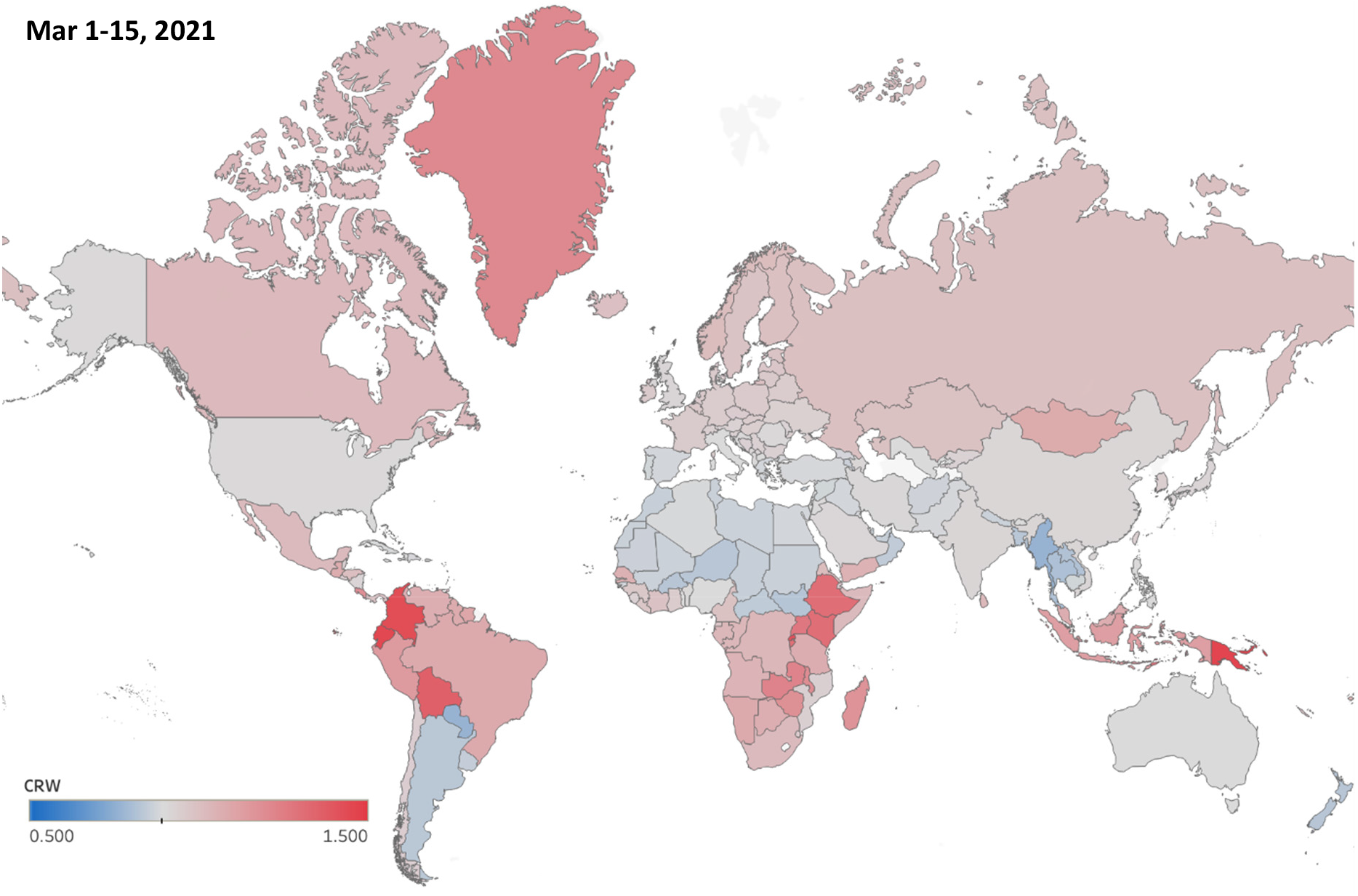
Projected Relative COVID-19 Risk Due to Weather (CRW), averaged for the first half of March 2021

## Footnotes

1 Similar to the main analysis, all models tested here use log(R_0_) as the outcome and include location fixed effects, location‐specific linear trends and day of the week effect. All models excluded days with new infections <1 and first 20 days since new infection exceeds 1 for the first time in each location unless specified otherwise.

## References

1. National Academies of Sciences Engineering and Medicine, “Rapid Expert Consultation on SARS-CoV-2 Survival in Relation to Temperature and Humidity and Potential for Seasonality for the COVID-19 Pandemic,” (Washington, DC, 2020).

2. J. Shaman, E. Goldstein, M. Lipsitch, Absolute Humidity and Pandemic Versus Epidemic Influenza. American Journal of Epidemiology 173, 127–135 (2010).

3. M. S. Nassar, M. A. Bakhrebah, S. A. Meo, M. S. Alsuabeyl, W. A. Zaher, Global seasonal occurrence of middle east respiratory syndrome coronavirus (MERS-CoV) infection. Eur Rev Med Pharmacol Sci 22, 3913–3918 (2018).

4. A. Altamimi, A. Ahmed, Climate factors and incidence of Middle East respiratory syndrome coronavirus. J Infect Public Health In press., (2019).

5. N. van Doremalen, T. Bushmaker, V. J. Munster, Stability of Middle East respiratory syndrome coronavirus (MERS-CoV) under different environmental conditions. Euro Surveill 18, (2013).

6. J. Yuan et al., A climatologic investigation of the SARS-CoV outbreak in Beijing, China. American Journal of Infection Control 34, 234–236 (2006).

7. L. M. Casanova, S. Jeon, W. A. Rutala, D. J. Weber, M. D. Sobsey, Effects of air temperature and relative humidity on coronavirus survival on surfaces. Appl Environ Microbiol 76, 2712–2717 (2010).

8. P. Jüni et al., Impact of climate and public health interventions on the COVID-19 pandemic: A prospective cohort study. Canadian Medical Association Journal, cmaj.200920 (2020).

9. M. M. Sajadi et al., Temperature, Humidity and Latitude Analysis to Predict Potential Spread and Seasonality for COVID-19. *Preprint*, (2020).

10. A. W. H. Chin et al., Stability of SARS-CoV-2 in different environmental conditions. The Lancet Microbe.

11. G. Kampf, D. Todt, S. Pfaender, E. Steinmann, Persistence of coronaviruses on inanimate surfaces and their inactivation with biocidal agents. Journal of Hospital Infection 104, 246–251 (2020).

12. N. van Doremalen et al., Aerosol and Surface Stability of SARS-CoV-2 as Compared with SARS-CoV-1. N Engl J Med, (2020).

13. L. Setti et al., SARS-Cov-2 RNA Found on Particulate Matter of Bergamo in Northern Italy: First Preliminary Evidence. *medRxiv*, 2020.2004.2015.20065995 (2020).

14. D. A. Glencross, T. R. Ho, N. Camina, C. M. Hawrylowicz, P. E. Pfeffer, Air pollution and its effects on the immune system. Free Radic Biol Med, (2020).

15. A. Notari, Temperature dependence of COVID-19 transmission. *medRxiv*, 2020.2003.2026.20044529 (2020).

16. G. F. Ficetola, D. Rubolini, Climate affects global patterns of COVID-19 early outbreak dynamics. *medRxiv*, 2020.2003.2023.20040501 (2020).

17. J. Bu et al., Analysis of meteorological conditions and prediction of epidemic trend of 2019-nCoV infection in 2020. *medRxiv*, 2020.2002.2013.20022715 (2020).

18. Q. Li et al., Early Transmission Dynamics in Wuhan, China, of Novel Coronavirus-Infected Pneumonia. N Engl J Med 382, 1199–1207 (2020).

19. C. Merow, M. C. Urban, Seasonality and uncertainty in COVID-19 growth rates. *medRxiv*, 2020.2004.2019.20071951 (2020).

20. W. Luo et al., The role of absolute humidity on transmission rates of the COVID-19 outbreak. *medRxiv*, (2020).

21. N. Islam, S. Shabnam, A. M. Erzurumluoglu, Temperature, humidity, and wind speed are associated with lower Covid-19 incidence. *medRxiv*, 2020.2003.2027.20045658 (2020).

22. B. Oliveiros, L. Caramelo, N. C. Ferreira, F. Caramelo, Role of temperature and humidity in the modulation of the doubling time of COVID-19 cases. *medRxiv*, 2020.2003.2005.20031872 (2020).

23. H. Qi et al., COVID-19 transmission in Mainland China is associated with temperature and humidity: A time-series analysis. Science of The Total Environment 728, 138778 (2020).

24. R. E. Baker, W. Yang, G. A. Vecchi, C. J. E. Metcalf, B. T. Grenfell, Susceptible supply limits the role of climate in the early SARS-CoV-2 pandemic. Science, eabc2535 (2020).

25. M. Lipsitch, in https://ccdd.hsph.harvard.edu/will-covid-19-go-away-on-its-own-in-warmerweather/. (2020).

26. A. Briz-Redon, A. Serrano-Aroca, A spatio-temporal analysis for exploring the effect of temperature on COVID-19 early evolution in Spain. Science of The Total Environment 728, 138811 (2020).

27. M. T. P. Coelho et al., Exponential phase of covid19 expansion is driven by airport connections. *medRxiv*, 2020.2004.2002.20050773 (2020).

28. K. M. O’Reilly et al., Effective transmission across the globe: the role of climate in COVID-19 mitigation strategies. Lancet Planet Health, (2020).

29. P. Shi et al., The impact of temperature and absolute humidity on the coronavirus disease 2019 (COVID-19) outbreak - evidence from China. *medRxiv*, 2020.2003.2022.20038919 (2020).

30. X.-J. Guo, H. Zhang, Y.-P. Zeng, Transmissibility of COVID-19 and its association with temperature and humidity. (2020).

31. Q. Bukhari, Y. Jameel, Will coronavirus pandemic diminish by summer? *Available at SSRN 3556998*, (2020).

32. S. A. Lauer et al., The Incubation Period of Coronavirus Disease 2019 (COVID-19) From Publicly Reported Confirmed Cases: Estimation and Application. Ann Intern Med, (2020).

33. N. M. Linton et al., Incubation Period and Other Epidemiological Characteristics of 2019 Novel Coronavirus Infections with Right Truncation: A Statistical Analysis of Publicly Available Case Data. J Clin Med 9, (2020).

34. E. Dong, H. Du, L. Gardner, An interactive web-based dashboard to track COVID-19 in real time. The Lancet Infectious Diseases.

35. W. J. Guan et al., Clinical Characteristics of Coronavirus Disease 2019 in China. N Engl J Med, (2020).

36. S. Zhang et al., Estimation of the reproductive number of novel coronavirus (COVID-19) and the probable outbreak size on the Diamond Princess cruise ship: A data-driven analysis. Int J Infect Dis 93, 201–204 (2020).

37. R. Li et al., Substantial undocumented infection facilitates the rapid dissemination of novel coronavirus (SARS-CoV2). Science, eabb3221 (2020).

## References

1. Q. Li et al., Early Transmission Dynamics in Wuhan, China, of Novel Coronavirus–Infected Pneumonia. New England Journal of Medicine, (2020).

2. S. A. Lauer et al., The Incubation Period of Coronavirus Disease 2019 (COVID-19) From Publicly Reported Confirmed Cases: Estimation and Application. Ann Intern Med, (2020).

3. W. J. Guan et al., Clinical Characteristics of Coronavirus Disease 2019 in China. N Engl J Med, (2020).

4. N. M. Linton et al., Incubation Period and Other Epidemiological Characteristics of 2019 Novel Coronavirus Infections with Right Truncation: A Statistical Analysis of Publicly Available Case Data. J Clin Med 9, (2020).

5. N. Popovich, in The New York Times. (New York, 2020).

6. P. D. Wibbens, W. Koo, A. M. McGahan, Projected COVID Infections, Deaths, and Local Social-Distancing Restrictions. Deaths, and Local Social-Distancing Restrictions (April 17, 2020), (2020).

